# Spatial pattern of all cause excess mortality in Swiss districts during the pandemic years 1890, 1918 and 2020

**DOI:** 10.1101/2024.02.13.24302701

**Authors:** Katarina L Matthes, Joël Floris, Aziza Merzouki, Christoph Junker, Rolf Weitkunat, Frank Rühli, Olivia Keiser, Kaspar Staub

## Abstract

**Background:** Every pandemic is embedded in specific spatial and temporal context. However, spatial patterns have almost always only been considered in the context of one individual pandemic. Until now, there has been limited consideration of spatial similarities or differences between pandemics.

**Objective:** The aim of this study was to examine spatial pattern of excess mortality in Swiss districts during the pandemic years 1890, 1918 and 2020. In addition, determinants that could explain the difference between districts were analyzed.

**Methods:** Excess mortality rate was estimated using a Bayesian spatial model for disease mapping. A robust linear regression was used to assess the association between ecological determinants and excess mortality.

**Results:** The highest excess mortality rate in all districts occurred during the 1918 pandemic, the lowest excess mortality rate was seen for the 1890 pandemic. Moreover, this analysis revealed heterogeneous spatial patterns of excess mortality in each pandemic year. Different socio-demographic determinants, in each pandemic, might have favored excess mortality. While the age composition, cultural and area-based socio-economic position differences and the proximity to France and Italy were the main determinants of excess mortality during the Covid-19 pandemic, the mobility, preexisting health issues (i.e. TB) or the remoteness location in the mountains played crucial roles during the historical pandemics.

**Contribution:** The analysis of spatial patterns in pandemics is important for public health interventions in future pandemics or outbreaks since it helps to identifying patterns of transmission. Identifying and understanding geographic hotspots informs precise interventions, aids in public health implementation, and contributes to tailored health policies for the region.

## Introduction

The most recent pandemic in Switzerland, Covid-19, was declared by the WHO in May 2023 as no longer a public health emergency of international concern^1^; nonetheless, in the last 150 years, this was not the first severe pandemic of respiratory pathogens in Switzerland. In the years 1918 and 1919, Switzerland was severely impacted from the influenza pandemic commonly referred as the “Spanish flu”, and 28 years prior, it was affected by the 1890 “Russian flu”. Since 1919, the severity of the pandemics and the numbers of deaths decreased in the 1957, 1969/70, 1977, 2003/2004 and 2009 pandemics. It is possible that this decline in the severity of pandemics as well as medical advances in the 20th century have led to an underestimation of the threat of a new pandemic^2^. Only with the onset of the Covid-19 pandemic in 2020^3^ has the scientific and public interest in past pandemic experiences increased sharply.

Each pandemic has a particular signature and specific characteristics – like a fingerprint – and it is embedded in a specific context in space and time ^4^. Experience with historical epidemics has shown how lessons from the past can be incorporated into a more effective responses^3^, especially in terms of strengthening society’s resilience and responsiveness to future crises. The aim is not to transfer ready-made recipes from history to the present. Rather, by contrasting and comparing past pandemics, the aim is to highlight the specificities of each pandemic and to offer the possibility of approaching current problems differently. Two studies from Switzerland have analyzed in detail the spatial spread of the “Spanish flu”^5^ and related public health interventions^6^ at the regional level (Canton of Bern), and have also made some comparison with the public health intervention during COVID-19.

Several recent publications examined the differences between Covid-19 and previous pandemics through various factors. For example, one study examined the differences in excess mortality over the past 140 years (including the 1890, 1918 and 1957 pandemics) between Switzerland, Spain, and Sweden. The study found geographical and pandemic differences in excess mortality, with Spain being the most affected during all three pandemics ^7^. In another study analyzing the different pandemic trajectories of the Covid-19 and 1918 pandemic in the United Kingdom, similarities were observe regarding the temporal trends^8^. Furthermore, an analysis of the socio-economic impact and economic repercussions of the 1918 and COVID-19 pandemics, conducted in both India and the USA^9,10^, underscored the significance of reflecting on historical events and incorporating the lessons learned from them. Another study conducted in the USA highlighted the impact of disparities in both urban and rural areas, as well as socio-economic differences, on mortality rates during both the 1918 influenza pandemic and the COVID-19 pandemic ^11^. Although numerous studies demonstrate the similarities and differences between the “Spanish flu” and Covid-19, very few link different pandemics through the context of spatial patterns. Some studies have indicated regional differences in morbidity and mortality during the different pandemics^12–21^. However, the potential reasons for these differences are multifaceted and contingent upon the particular characteristics of each pandemic. For instance, factors such as the availability of railroad connections, the incidence of tuberculosis, pre-existing regional conditions, the density of factories and their workforce in a community, as well as factors related to poverty and socioeconomic status, have exerted significant influence on the pandemics of 1890 and 1918^12–17^. As for the Covid-19 pandemic, socioeconomic status, poverty, and inequalities in the healthcare system continue to play an important role^17–22^.

Spatial patterns have almost always only been considered in the context of a single pandemic where the consideration of spatial similarities or differences between pandemics has been limited. Beyond factors such as age, sex, and socio-economic status, spatial factors play a crucial role in enhancing our comprehension of pandemics and their consequences. Furthermore, spatial distribution helps to recognize transmission patterns that are caused, for example, by population density, transport networks, proximity to infected individuals, healthcare infrastructure, and more. In addition, identifying and understanding geographic hotspots can enable targeted interventions, allocation of resources and public health measures, as well as help with the development of health policies and interventions tailored to the specific needs of a region.

This study, therefore, examines and compares the spatial differences in excess mortality among Swiss districts during the last three significant pandemics in Switzerland. The pandemic of 1890, also known as the “Russian flu”, was the first pandemic in the industrial world with a well-developed rail network^16^.It reached its peak in Switzerland in January 1890, followed by only a few smaller waves persisting until 1894. Approximately 1 million global fatalities were attributed to the “Russian flu” ^23^, while Switzerland had reported approximately 3000 deaths^24^ with an associated excess mortality rate of 6%^7^. Several hypotheses exist regarding the etiology of the 1890 pandemic, with one positing an influenza virus (H3N8) ^25^. However, contemporary studies do not dismiss the possibility of a coronavirus (HCoV-OC43) due to observed clinical symptom similarities with Covid-19^26^. The initial wave of the 1918 pandemic (H1N1 influenza virus), commonly referred to as the “Spanish flu,” impacted Switzerland in late June 1918. This was succeeded by a longer and more severe second wave spanning from October 1918 to the early months of 1919, followed by a subsequent wave at the onset of 1920. Over 50 million worldwide lives were taken during this pandemic ^27^ of which 25000 deaths^28^ and an excess mortality of 50%^7^ were approximately accounted for Switzerland. The most recent pandemic, COVID-19 (SARS-CoV-2), resulted in more than three million deaths worldwide in 2020 ^29^, where7610 deaths^30^ and an excess mortality of 12.5%^7^ were recorded in Switzerland. In each of the three pandemics, particular emphasis is placed on the first year of the outbreak, wherein a substantial portion of the population remained largely susceptible, lacking immunity or vaccination.

Switzerland is particularly pertinent country to explore spatial variations at the district level, as political decisions, general healthcare practices, and pandemic mitigation strategies are formulated at the cantonal level or even lower administrative tiers ^6,31^. Additionally, Switzerland is characterized by its multilingualism and three main language regions (German, French, and Italian) and therefore exhibits distinct cultural diversities influenced by neighboring countries (refer to Supplementary Figure 1). These cultural differences also lead to regional differences in health status and mortality^32,33^. Moreover, the absence of territorial changes in Switzerland between pandemics, coupled with its non-involvement in the First World War, ensures good data quality.

The aim of this study is to analysis the total, age and sex specific spatial excess mortalities of three pandemics and associate the excess mortality to various ecological determinants.

## Data

### Death counts

The annually number of all death causes from 1882 to 1920 stratified by district, sex and age group from the “Annual population movements of the Federal Statistical Office” were obtained by the Swiss Federal Statistical Office (FSO) and converted into machine readable files by the study team.

The aggregated annual number of deaths for each district, sex and age group for 2016-2020 was delivered to the study team by the Swiss Federal Statistical Office^34^. This historical all-cause death data is deemed of high quality and comparable over time^35^. Nonetheless, concerns related to the historical mortality data primarily evolve around the cause of the reported deaths rather than the overall count of the death data. The specific regulations of the Swiss government on the official reporting of a death outlined the precise requirements regarding when and how such reports were to be made^36^. Therefore, completeness, at the close of the 19th century, may have only been affected by under-registration of deaths occurring in the initial days or weeks of life or very advanced ages and due to emigration^37^.

### Population

Census data of the years 1880, 1888, 1900, 1910 and 1920 for each district and sex was provided by the FSO^38^. For the years 1888 and 1910, additional population data were available for age groups in 5-year intervals (i.e., 0-4, 5-9, 10-14, etc.). Annual population estimates by sex and age was linearly interpolated for each district using census data of the years 1880, 1888, 1900, 1910, and 1920. Afterwards, the age distribution of the years 1888 and 1910 for each district was calculated. These distributions were then used to estimate the population of each age group for the remaining years, where the distribution from 1888 was used to estimate age-specific population estimates from 1880 to 1900 and the distribution from 1910 was used for the years 1901 to 1920. The population data for 2016 – 2020 was provided by the FSO ^39^.

## Methods

### Districts

The number of districts in Switzerland has changed over the years. In 1882, a total of 183 districts existed due to mergers over the years; however, by 2020, only 143 districts were left. For the spatial comparability of pandemics, the number of districts from 2020 were used, while the districts that were separate in 1890 and 1918 were combined (40 districts in 1918 and 1890 have been merged). For the districts that were combined, the older districts were used (12 districts in 2020 were merged to 6 districts). As for the small canton of Schaffhausen, the mortality data of its six districts was only accessible in aggregated cantonal form. Therefore, for our study, these six districts were merged into one cantonal district called “Schaffhausen”. After the harmonization between the districts of 1890, 1918 and 2020 the total number of comparable districts resulted in 130 identical districts for all time periods.

### Excess mortality

The annual excess mortality rate for each of the 130 districts for each year was estimated using a Bayesian spatial model for disease mapping under the scenario of no pandemic, where death and population data from the 4 years prior to the pandemic was utilized. Because the population in some small districts is very small with not many or any deaths per year, we used a zero inflated negative binomial model for the number of deaths, while the population was considered as an offset. To incorporate the spatial structure into the model, the BYM (Besag-York-Molliè) model ^40^ was used. Additionally, the year was included as an independent and identically distributed Gaussian random effect. After model fitting, we drew 1000 samples from the posterior distribution (i.e. the expected number of deaths) and calculated the median and 95% credible intervals (CrIs). The respective excess rates were calculated by subtracting the expected values from the observed ones. The excess mortality rates were displayed in percent. The excess mortality rates of the respective pandemic years and districts were shown on a choropleth map using “Jenks natural breaks” to define ranges. Estimates of excess mortality were made separately for each pandemic, each sex and the age groups 0-69 years and ≥ 70 years old. As deaths throughout the years and districts were not reported in all younger age groups and therefore excess mortality could not be precisely estimated, we limited the analysis to two age groups. Finer age groups were only considered in an additional analysis (0-19 years, 20-29 years, 30-39 years, 40-69 years, ≥70 years) for the 1918 pandemic, where the younger population experienced more deaths. Differences in excess mortality within a pandemic and between sex and age groups, respectively, were calculated using linear regression, with excess mortality as the outcome and sex or age as the explanatory variable.

### Local spatial statistic G

Local spatial statistic G was used to cluster the districts with higher or lower excess mortality rates. The G statistic represents z-values. Higher z-values indicate greater intensity of clustering and the direction (positive (red color) or negative (blue color)) indicates a cluster of high or low excess mortality rate. The results for each pandemic are displayed on a choropleth map.

### Determinants

Ecological determinants that could influence excess mortality in the individual districts were collected. In all three pandemics, we calculated the population density for each district and defined population density groups based on the median. For these pandemics, school children are often seen as drivers of a pandemic^41^. Unfortunately, since information regarding the opening and closing of schools during the 1890 and 1918 pandemic for each district was unavailable, a proxy was used. As a proxy, we have calculated the proportion of school children per district where higher proportions were assumed to lead to higher incidence and therefore a higher excess mortality. Additionally, it is known that Covid-19 mainly affected the older population and the 1918 pandemic the younger population, which is why the proportion of ≥ 70-year-olds per district was also considered. In addition, the proportion of men living in a district was used as a determinant of excess mortality. Furthermore, each district was defined as an urban or rural district, assuming that urban areas are more affected ^42^. The socio-economic position (SEP) is known to be correlated to health and mortality ^21,43–45^. To reflect it, the Gross Domestic Product (GDP) ^46^ per capita for 1890 and 1918 was used. As for the 2020 pandemic, since information on GDP per district was unavailable, we used the Swiss neighborhood index of socioeconomic position (Swiss-SEP), as described elsewhere^47,48^. The Swiss-SEP provides an even more precise indicator then the GDP to display health and mortality differences in districts. For the pandemics of 1890 and 1918, we collected additional factors that could be related to excess mortality. As an indicator of access to healthcare, the number of hospitals in each district was used ^49,50^ and classified in two groups, namely no hospital and at least one hospital. Furthermore, we calculated the proportion of child mortality (0-4 years old) one year before the pandemic as a proxy for the general health status in a district. Since the pandemic of 1890 took place during the industrialization and urbanization of Switzerland and the development of railway networks, with the modernization of transport and the commuting of workers, infection through travel by those who became ill was encouraged and the influenza virus spread rapidly, in particular along the train routes ^17,24,51^. Therefore, the number of Swiss railway stations of the year 1900 ^52^ per 1’000 inhabitants was used as a proxy of mobility for the inhabitants of a district for 1890 and 1918 pandemics. Pulmonary tuberculosis (TB) is considered a risk factor for influenza mortality and higher excess mortality^53,54^. Moreover, at the same time as the pandemics of 1890 and 1918, TB also reached a mortality peak in Switzerland^55^, which may have contributed to the high excess mortality. Therefore, the number of deaths due to TB were also considered as determinants explaining excess mortality. However, since data on the sum of TB deaths were only available for the years 1885-1890, we calculated, using the annual population data, the mean TB mortality per year. For the 1918 pandemic, the sum of TB deaths from 1911-1920 was available, allowing us to calculate the mean TB mortality per year. Using the ecological determinants for each district, the relationship between each determinant and excess mortality was examined in an exploratory analysis using robust linear regression (to overcome the issue of outliers and extreme values). The regression coefficients and the 95% confidence intervals are displayed in a figure.

All statistical analyses were performed using R Version 4.2.2.^56^. R-INLA ^57,58^was used to estimate the expected mortality. The R packages “sf”^59^, “spdep” ^60^, and “tmap” ^61^ were used to calculate the spatial clusters and to present the choropleth maps. “MASS” ^62^ was used for the robust linear regression and “tidyverse” ^63^was used to process the data and create the remaining figures.

## Results

The overall mortality in Switzerland was 208.1 per 10’000 inhabitants in the year 1890, 194.2 in 1918, and 87.9 in 2020. Although the overall mortality level in 1890 was higher than in 1918 across Switzerland, the Supplementary Figure 2 shows that in 1918, some districts experienced a higher mortality rate than in 1890, particularly in the canton of Valais in Southern Switzerland. Comparing the excess mortality of all three pandemic years (Figure 1), it is clear that the highest overall excess mortality rate in all districts occurred in 1918 when the “Spanish flu” occurred, while the lowest excess mortality rate was in 1890 when the “Russian flu” occurred. In 1918, 97% of the districts had an excess mortality, whereas in 1890 the observed excess mortality was shown in around 15 % of the districts. In 2020, during Covid-19, excess mortality was observed in 75% of the districts. In the Supplementary Tables 1-3, the observed, expected values and CrIs of each the respective district and pandemic year are displayed. Considering the local G statistics, a cluster of higher excess mortality appeared in the canton of Ticino (the Italian-speaking South of Switzerland) and in the canton of Aargau (Northern Midland of Switzerland), in 1890. In 1918, the canton of Valais, in the South, showed a clear clustering of higher excess mortality compared to the rest of Switzerland. It is important to highlight that the blue regions don’t signify low excess mortality in other districts. Excess mortality remained high across all districts, with the canton of Valais experiencing even higher rates. As for the 2020 pandemic, a clustering of higher excess mortality, especially in the French- and Italian-speaking regions of Switzerland, are shown.

**Figure 1:**
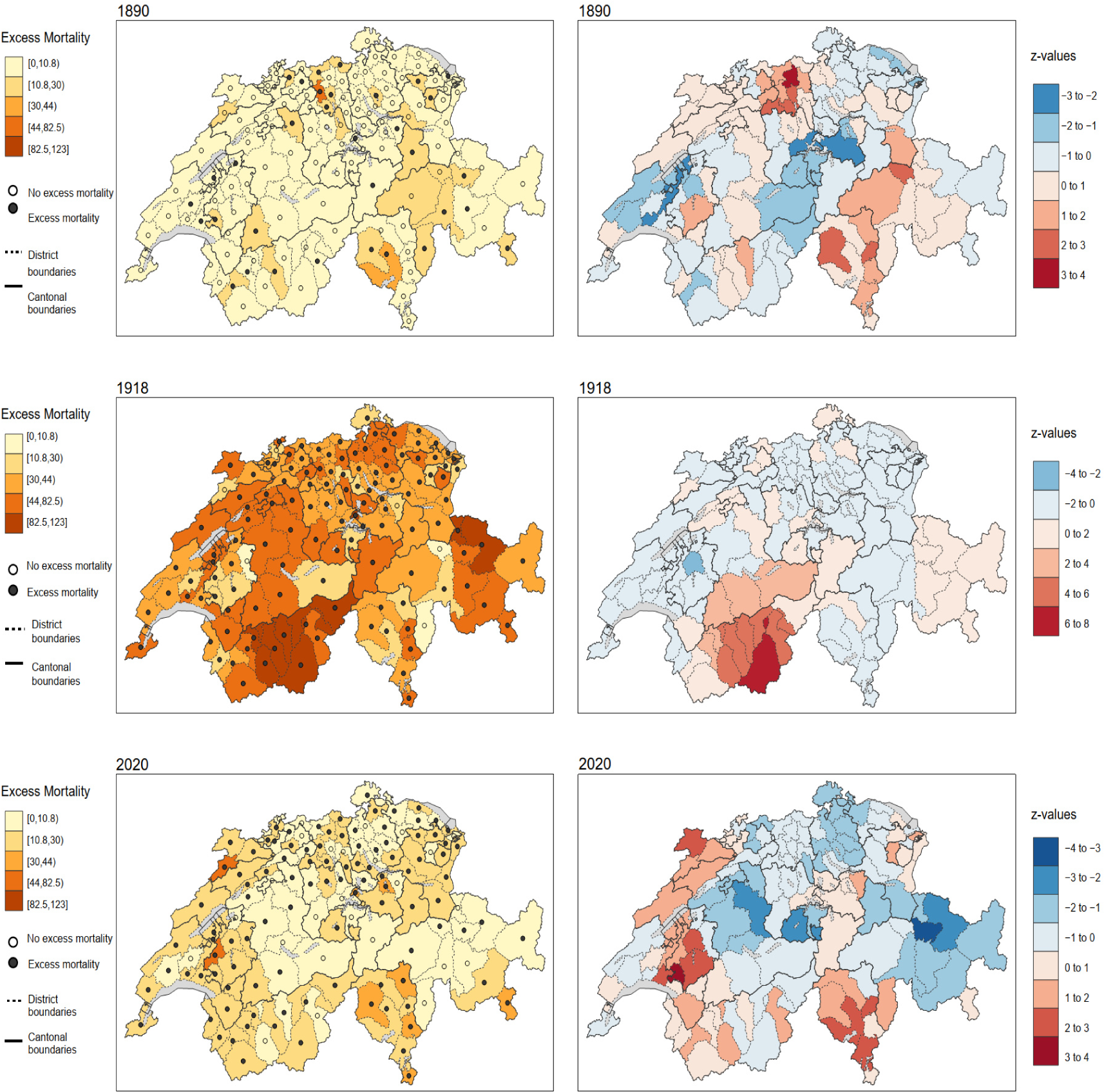
Excess mortality of each pandemic year (left) and results of the local spatial statistic G (right). Higher z-values indicate greater intensity of clustering and the direction (positive (red color) or negative (blue color)) indicates a cluster of high or low excess mortality rate.

In Figure 2, the spatial differences between the sexes is shown for each pandemic year. In 1890, the excess mortality rate was slightly higher among females (-0.2 % difference [CI: -2.4 to 2.0 %]). Females showed a higher excess mortality than men, especially in Ticino and Eastern Switzerland. In 1918, the excess mortality rate was high for both sexes, but lower for females than for males (9.7% difference [CI: 3.9 to 15.5%]). However, both sexes were highly affected in Valais whereas fewer females were affected in Central Switzerland. In 2020 the excess mortality rate was clearly higher for males compared to females (4.0% difference [CI:0.89 to 7.2%]), especially in the French- and Italian-speaking regions. In Central Switzerland, the differences between the sexes were less prominent.

**Figure 2:**
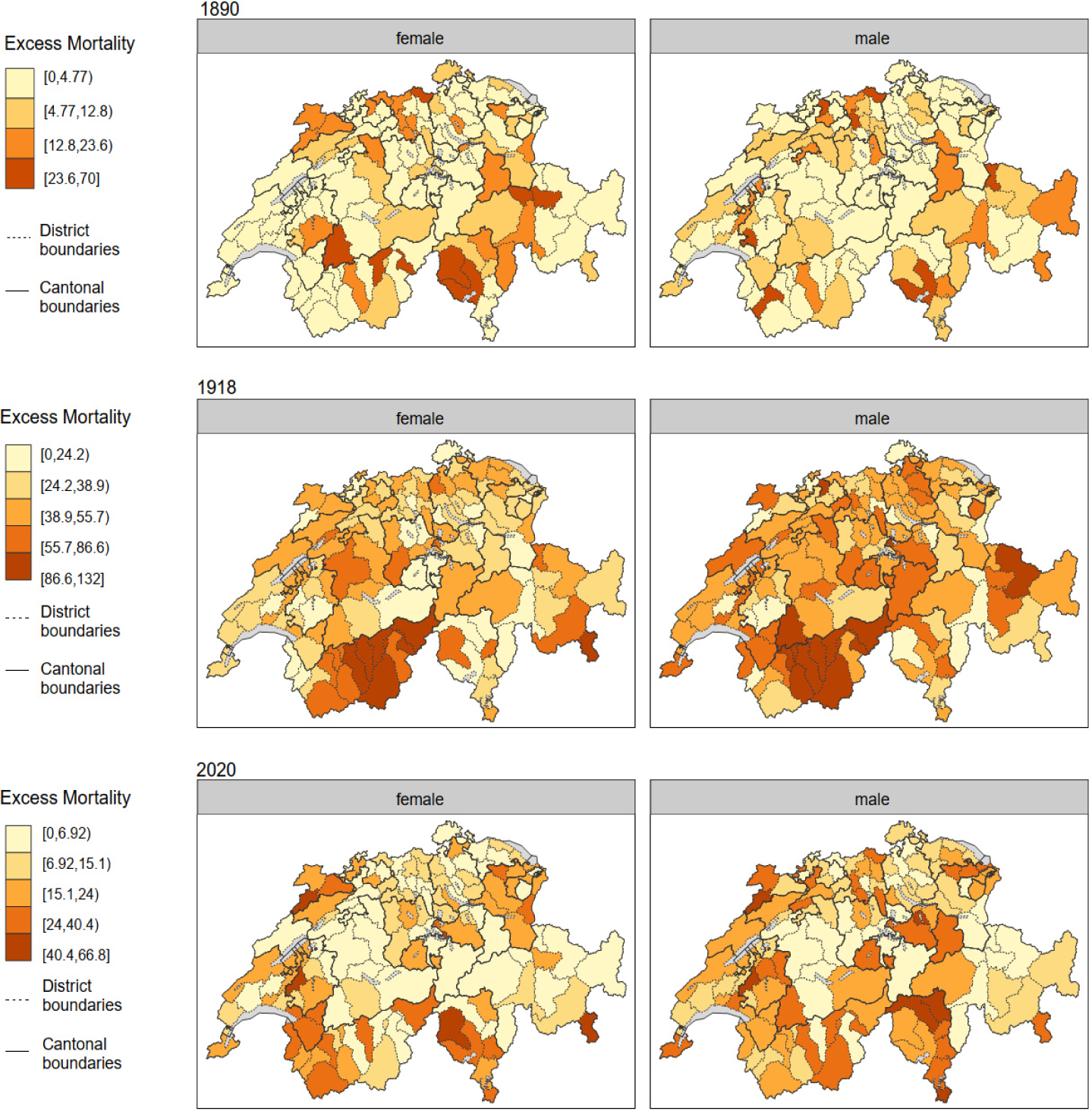
Sex specific excess mortality of each pandemic year.

Figure 3 displays differences by age groups for each pandemic year. In 1890 excess mortality rate was higher for people aged ≥70 age (3.0 % difference [CI: 0.4 to 5.7%]). The greatest differences between age groups can be seen in the north of German-speaking Switzerland and Ticino. In 1918, excess mortality was much higher among those aged 0 to 69 years (50.6% difference [CI: 45.8 to 55.3]). However, in Valais, excess mortality was also high for those aged ≥ 70 years. In 2020, the excess mortality rate was higher among those aged ≥70 (5.4% difference (CI: 2.4 to 8.3%)). A high excess mortality in those ≥ 70 years old was particularly noticeable in Ticino and in the French-speaking parts of Switzerland. Since the excess mortality rate in 1918 was particularly high in people under 40 years, we also investigated finer-grained age groups for the “Spanish flu” (Figure 4). Based on this figure, it can be seen that the age group of the 20 to 29 years-old was particularly hit hard, followed by the age group of 30 to 39 years old. However, the highest excess mortality for each age group was again observed in the districts of the Canton of Valais. Among those aged 20 to 29 years, during the 1918 pandemic (see Supplementary Figure 4), excess mortality rate was much higher among males than females (119. 0% difference [CI: 93.4 to 144.6%]), but in some districts of Valais the excess mortality of females was similar or even higher than for males.

**Figure 3:**
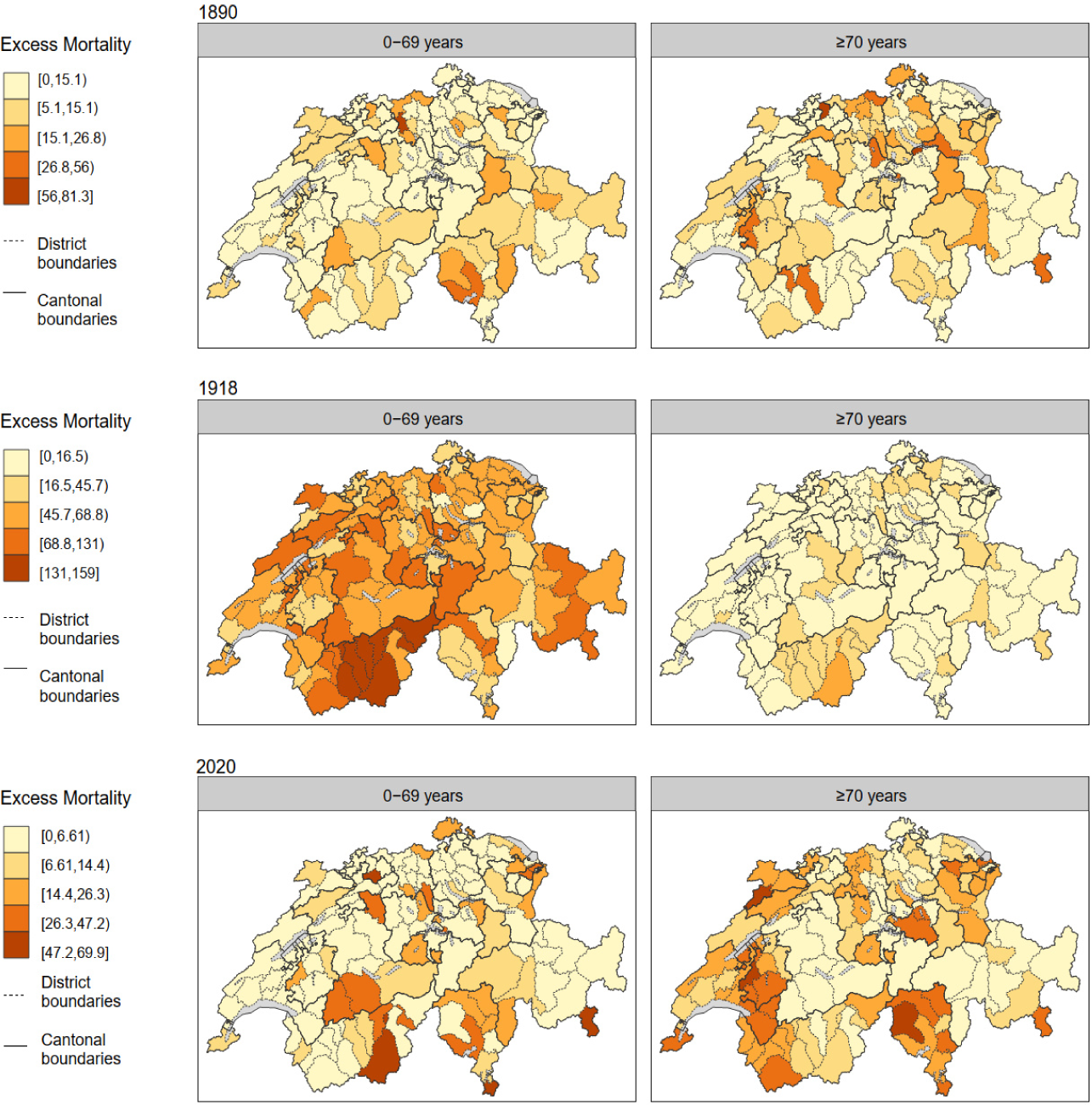
Age specific excess mortality of each pandemic year.

**Figure 4:**
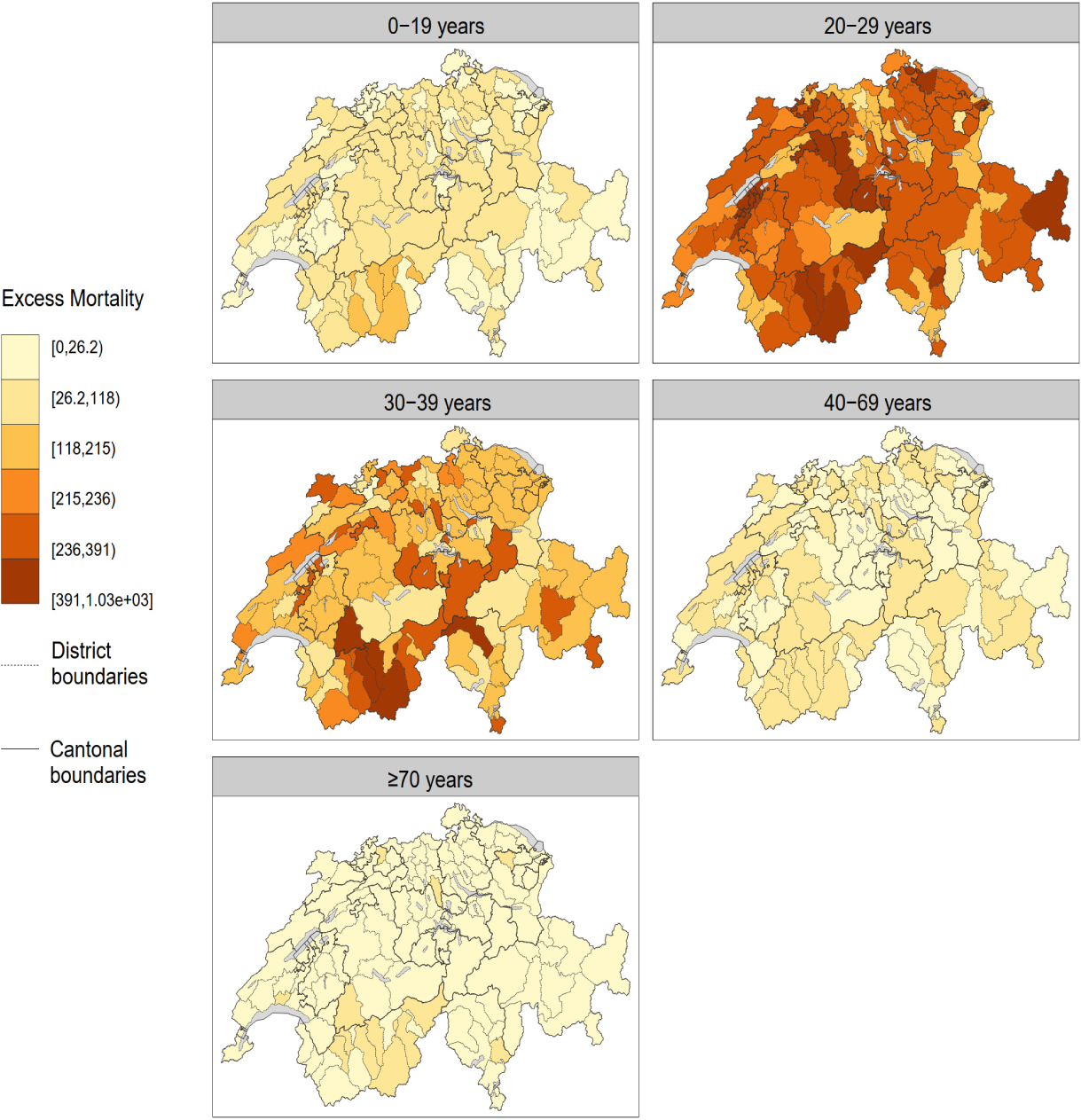
Age specific excess mortality of the 1918 pandemic year.

The results of the robust linear regression are shown in Figure 5. In 2020, a higher Swiss-SEP is associated with a lower excess mortality (-11.75 % [CI: -19.57 to -3.93]), meaning that lower area-based SEP-districts were hit harder. In 1890, a higher population density was associated with lower excess mortality (-3.64 % [CI: -7.34 to -0.06], and a higher tuberculosis mortality was associated with a higher excess mortality (3.24 % [CI: 0.93 to 5.54]). The proportion of people age ≥ 70 years was associated with lower excess mortality, in 1918 (-2.59 % [CI: -4.83 to -0.36], and a higher mortality in 1890 (2.63 % [CI: 0.66 to 4.60]. The proportion of men in each district was associated with a lower excess mortality in 1890 (-24.12 % [CI-37.02 to -11.22]) and a higher in 1918 (25.54 [CI: 3.07 to 48]). The results of the regression models are shown in Supplementary Figure 5-24 and Table 4, where the distribution of these determinants through pandemic years and language regions can be observed.

**Figure 5:**
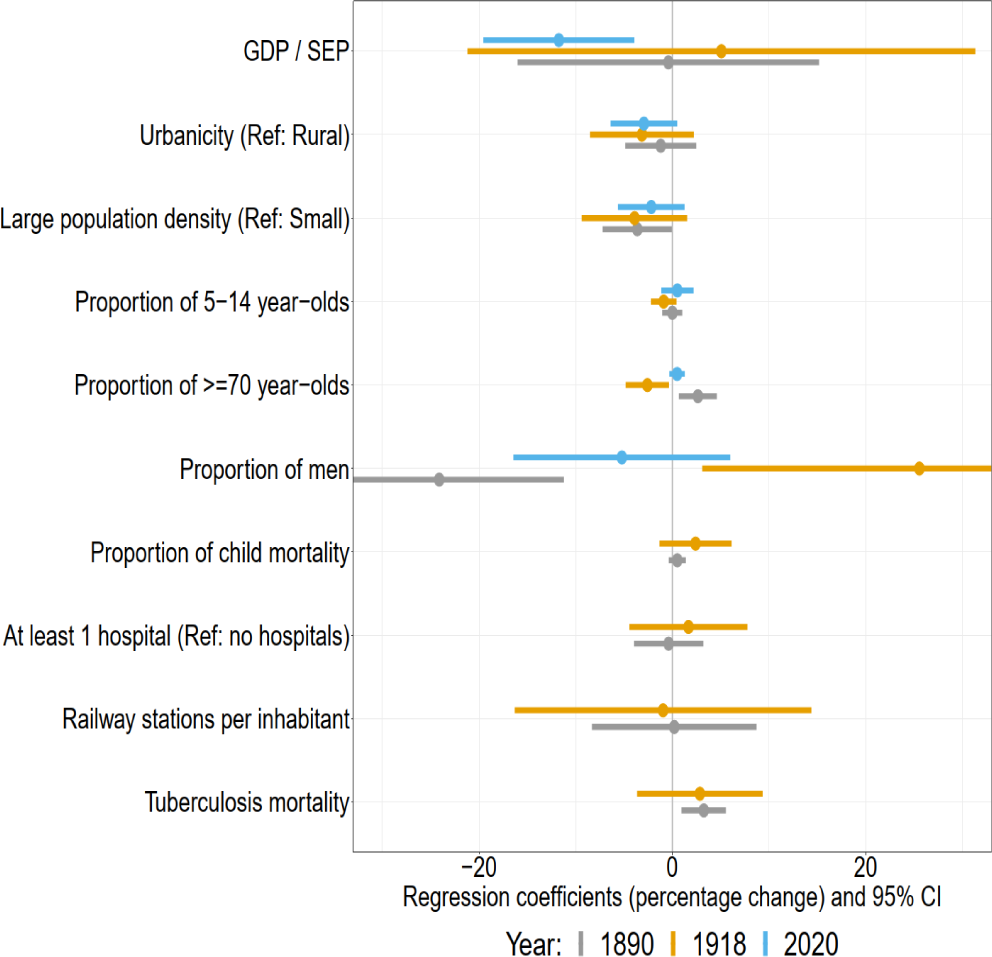
Percentage changes (regression coefficients) in excess mortality and 95 % confidence interval (CI) of the robust linear regression. Each determinate is showing one model (univariable)

## Discussion

We conducted a spatial analysis of the three last major pandemics in Switzerland by estimating the annual all-cause excess mortality per district and by sex and age. This analysis revealed heterogeneous spatial patterns of excess mortality in each pandemic year. Different socio-demographic determinants, in each pandemic year, might have favored excess mortality. In 2020, the SEP played a decisive role in excess mortality where a low SEP led to higher excess mortality. In the historical pandemics, previous illness with tuberculosis was an important factor for higher excess mortality (which could also be an indicator of poverty), as was age where during the 1918 pandemic, districts with a younger population were particularly affected whereas districts with an older population were more affected during the 1890 pandemic.

During the 1890 pandemic year, the districts were similarly affected with a relatively low all-cause excess mortality, but with significant age differences. Districts with people aged ≥70 years were slightly more affected as well as districts with a higher proportion of women. Our results confirm a study using annual flu mortality data from Connecticut, USA, which have also shown only small differences in age groups and a slightly higher death rate in females^64^. Using summarized weekly flu death data of the 15 largest city from Schmid (Statistical reports on influenza data for the years 1889-1894 from the Swiss Health Office in 1895)^24^, Valtat et al. found that, in Switzerland, a higher flu mortality occurred in the ≥60 years age group^65^. Ramiro et al., using daily and weekly mortality data from Madrid, showed a higher excess mortality for age groups ≥50 years^66^. Although the severity of the 1890 pandemic was relatively mild and the case-fatality rate rather low (ca. 0.1% to 0.28%)^16^, excess mortality was observed globally ^16,64–67^, especially excess death for deaths from pneumonia and respiratory diseases. The 1890 pandemic was so widespread (in only 4 months the virus was present worldwide ^16^), that even with a relatively low case fatality rate, the increasing number of deaths lead to an excess mortality^64^. In Switzerland, the flu epidemic probably only lasted a short time from December 1889 to February 1890^24^. There was apparently another stronger wave in the winter of 1893 to 1894 ^24^. When comparing our spatial excess mortality maps with the regional flu mortality data of Schmid ^24^, both regional maps were relatively consistent. In both studies the highest mortality was observed in North Switzerland, Ticino and Valais. Le Goff ^51^ investigated the spread of the flu in Switzerland using the data of Schmid^24^, information of railway stations, and distances between communities. This study showed that the spread first took place through places that belonged to the same railroad company (there were 5 different ones), and that these communities carried the infection further to their neighboring municipalities. According to this study, the density of railway stations was particularly high in Northern Switzerland, where the mortality rate was among the highest in our study and also in Schmid’s study.

The 1918 pandemic was the most severe in all districts of Switzerland with a case fatality rate 10 times higher compared to the 1890 pandemic ^16^. The canton of Valais appears to have the highest excess mortality rate in Switzerland, which was also confirmed by a study investigating the “Spanish flu” in canton of Valais ^68^. The reasons for the highest mortality in Switzerland, given by the author, were, among others, the living conditions that prevailed in Valais in 1918. Valais is a mountainous canton with many valleys that were still isolated at that time. Poor hygiene, unsanitary housing, elevated alcohol consumption, poverty, as well as malnutrition had adverse consequences. This situation was exacerbated by the lack of health infrastructure in Valais at that time ^68^. However, conditions were similar in other Alpine regions at this time, in which the excess mortality rate was not as high as in the canton of Valais. Excess mortality in Italian-speaking Switzerland was lower compared to the other parts of Switzerland, although Italy was one of the most affected countries^69^. One reason could be that Ticino was affected later than the rest of Switzerland as there was no summer wave here and the strong second wave only began at the end of 1918 to beginning of 1919. However, if we look at the excess mortality of the Ticino districts in 1919 (Supplementary Figure 3), the excess mortality in Ticino was no higher than in 1918. In 1918, more public health measures were taken to combat the pandemic than in 1890. In the first wave (summer 1918), measures were taken at the cantonal level, whereas at the beginning of the second wave (autumn 1918), the cantonal authorities in the Cantons of Zurich and Bern transferred responsibility for the implementation of measures to the municipal authorities for 1-3 weeks^6^.Nevertheless, no detailed information was available for each district or municipality. Various hypothesis regarding the extreme mortality in young adults exist. One of them hypothesizes is that the elderly were previously exposed (before the 1890 pandemic) to a similar type of virus as the one from the 1918 pandemic and were therefore protected through cross-protection)^70,71^. Early exposure in person, or even in-utero, to the 1890 influenza virus, which was not the same type of virus as the 1918 pandemic, could also have led to an increased susceptibility to a severe course of the 1918 pandemic^72,73^. Another hypothesis is that the young adults were prone to an immunopathogenic reaction (cytokine storm) after H1N1 infection (1918 pandemic) ^74^ or that the high mortality is also due to the simultaneous high incidence of TB ^75^. However, the increased mortality of young adults is probably due to multiple factors. A few studies have analyzed spatial patterns and have shown that a higher proportion of children were associated with higher excess mortality ^14,15^, which was not the case in our study. A study from Portugal has found associations between excess mortality and geographic or socio-demographic factors, but these differed by pandemic wave ^76^. A Dutch study has shown that spatial cluster of excess mortality especially occurred in the second wave ^13^. Unfortunately, our study did not allow us to evaluate excess mortality by pandemic waves but only by pandemic years.

Our study shows, that Covid-19, the most recent pandemic, has lower excess mortality than the 1918 pandemic, but higher than the 1890 pandemic. Our findings are consistent with those in earlier publications examining sex and age differences of the Covid-19 pandemic ^77–80^. Konstantinoudis et al.^81^ have compared the Covid-19 excess mortality of different larger regions of Europe and spatially determined excess mortality. The results for Switzerland confirm the findings of this study while also showing a lower excess mortality in eastern Switzerland (Grisons) and a higher mortality in the Ticino (Italian-speaking Switzerland) and in the French-speaking part of Switzerland. Additionally, it showed how severely northern Italy was affected by the Covid-19 pandemic ^81^. The high excess mortality in Ticino could therefore be explained by its close geographical position with Northern Italy and by the fact that Ticino was the first canton report a Covid-19 case in Switzerland. Furthermore, it has been shown that the eastern part of France (bordering Germany and Switzerland) had the highest excess mortality in France^82,83^, which could also lead to the higher excess mortality in the French-speaking part of Switzerland. The canton of Geneva in particular was hit very early and hard by Covid-19 ^84^.Pleninger et al. ^31^ adapted the Covid-19 stringency index for Switzerland and compared the Swiss cantons regarding the measurements applied during Covid-19. From March to June 2020, the measures were the same throughout Switzerland. After that, it becomes apparent that cantons that were hit hard in the first wave in spring 2020 (Italian and French-speaking parts of Switzerland) took stronger measures from July 2020 onwards, reflecting the mortality rate. As annual data could only be used in our study, regional monthly differences or differences between the waves could not be analyzed. Nonetheless, according to a study, the Italian and French-speaking districts showed a higher excess mortality in the first wave (spring 2020) than the German-speaking districts^85^.

TB and excess mortality were significantly associated in 1890 and showed a positive trend in 1918 (although not significant). These results confirm the results of Zürcher et al. showing a high peak of TB and influenza especially in 1890 ^55^. However, we have no data on influenza deaths in our study, and can therefore not distinguish whether the excess mortality was more likely due to influenza or TB. Nevertheless, the cause of death might also be misclassified, as TB and influenza deaths were often confused^55^. Therefore, it is likely that a combination of both diseases led to increased excess mortality in 1890. TB showed a higher mortality for the ≤ 60 years old, and influenza a higher mortality in older adults ^55^. The proportion of people ≥ 70 years in the district is significantly associated with excess mortality, which could indicate excess mortality due to influenza in these districts. In 1918, in addition to TB, mortality from influenza was also high in young people. TB might be a risk factor for influenza mortality, but cannot be the sole reason for a high excess mortality in young adults^86^. Although controversial, a smaller population density was significantly associated with a higher excess mortality in 1890, which was similarly shown in a Dutch spatial study of the 1918 pandemic. This suggests that factors other than population density, such as poorer living conditions or malnutrition in rural areas, could play a role in excess mortality in historical pandemics. These factors could also explain the lower excess mortality in urban areas in our study. In 1890 and 1918 the number of hospitals did not seem to be associated with excess mortality, which lines up with a study from Denmark showing that access to medical care were not associated with increased mortality ^42^. In our study, however, we could not investigate access to medical doctors or the effective distance to medical care. The association between a lower area-based SEP and more severe Covid-19 courses have, been investigated in several studies ^87–89^ confirming our results. For instance, Riou et al. ^21^ showed that people in Switzerland with a lower area-based SEP were less likely to be tested for Covid-19, had a higher probability to test positive, develop a severe course of the disease and a higher probability of death compared to people with a higher SEP. In our study, the ecological determinants at the district level may be too imprecise to reveal more precise associations between the determinants and excess mortality, where analyses at the municipality level might be more precise. Another paper, which only analyzed morbidity data of the Canton of Bern (Switzerland) in the respective municipalities, could show associations of some determinants and incidence rates during the 1918 pandemic ^5^. For example, municipalities with at least one railway station had a higher risk of higher incidence rate during the first wave of the 1918 pandemic, but not during the second or third waves. This could confirm the hypothesis that in historical pandemics, cities with a higher mobility were the first to be affected. In addition, a higher proportion of industry occupational sector leads also to a higher incidence rate ^5^. Unfortunately, we did not have information about the number of factories for each district in Switzerland, in this study.

The calculation of excess mortality has some limitations, especially for the 1890 and 1918 pandemic. The underlying population was estimated for the historical pandemic years, and for 1890 interpolated population data between 1888 and 1900 was used. Since the 1890 pandemic was not as deadly as the 1918 pandemic and the census of 1888 is close to 1890, the estimated population at risk in 1890 is probably slightly biased. However, for 1918, the interpolated data from the 1910 and 1920 census was used where the age distribution was only available for 1910. In the years between 1910 and 1920, there were major changes in the population pyramid, first due to World War I (but with only few war-related deaths) and then due to the pandemic itself. This means that the estimated population at risk in 1918 is more likely biased, but because Switzerland did not actively take part in World War I, the bias is probably smaller than in other countries. In addition, migration is not considered in the estimation of the population. Despite the limitation of the population estimations, we believe that an interpretation of the estimated excess mortality is still plausible, since we compared in the districts. But caution is still required in the exact numerical interpretation of the excess mortality of the 1890 and 1918 pandemics. Furthermore, we only estimate annual excess mortality and not monthly data. But, the pandemic of 1890 was mainly severe in January, and in 1918, the second half of the year was affected by the pandemic. Similarly, in 2020, not all months were equally affected by Covid-19. This most likely results in a lower annual excess mortality rate than in the respective months when the pandemic occurred.

## Conclusion

The spatial patterns of excess mortality differ between the pandemic. The reasons are manifold and not the same depending on the pandemic. While the age composition, cultural and area-based SEP differences and the proximity to France and Italy were the main determinants of excess mortality during the Covid-19 pandemic, the mobility, preexisting health issues (i.e. TB) or the remoteness location in the mountains played crucial roles during the historical pandemics. Analyzing spatial distributions in pronounced multicultural countries like Switzerland is important in determining and establishing public health interventions for future pandemics and outbreaks and in identifying patterns of transmission. In addition, the identification and comprehension of geographic hotspots can empower precise interventions, efficient resource allocation, implementation of public health measures, and contribute to the formulation of health policies and interventions tailored to the specific needs of a region. Although our study has already show spatial differences in excess mortality, additional studies at the municipality level, macro level or studies with individual data would be helpful and important to better understand the various factors that lead to excess mortality.

## Author statements

## Ethics committee approval

Ethics approval was not required for the reuse of these aggregated and publicly available Federal authority data.

## Data and code

The data and code underlying this manuscript are publicly available via zenodo and GitHub: https://github.com/KaMatthes/UZH_UniGe_Past_Pandemics

## Data Availability

All data produced are available online at https://github.com/KaMatthes/UZH_UniGe_Past_Pandemics

## Acknowledgements

We would like to thank the Swiss Federal Statistical Office for providing the aggregated modern data for 2016-2020, the Institute for Social and Preventive Medicine at the University of Bern (Marcel Zwahlen, Radoslaw Panczak, Claudia Berlin) for providing the modern Swiss-SEP information on the district level, Konstantin Büchel and Stephan Kyburz for providing historical information on railway stations, and Julia Simola und Inga Birkhäuser for digitizing the historical data. We thank Lucas Bianchi, Daniel C. P. Câmara, Flavio Coelho and Sabina Rodriguez Velásquez for helpful comments and support.

## Supplement

**Supplement Figure 1:**
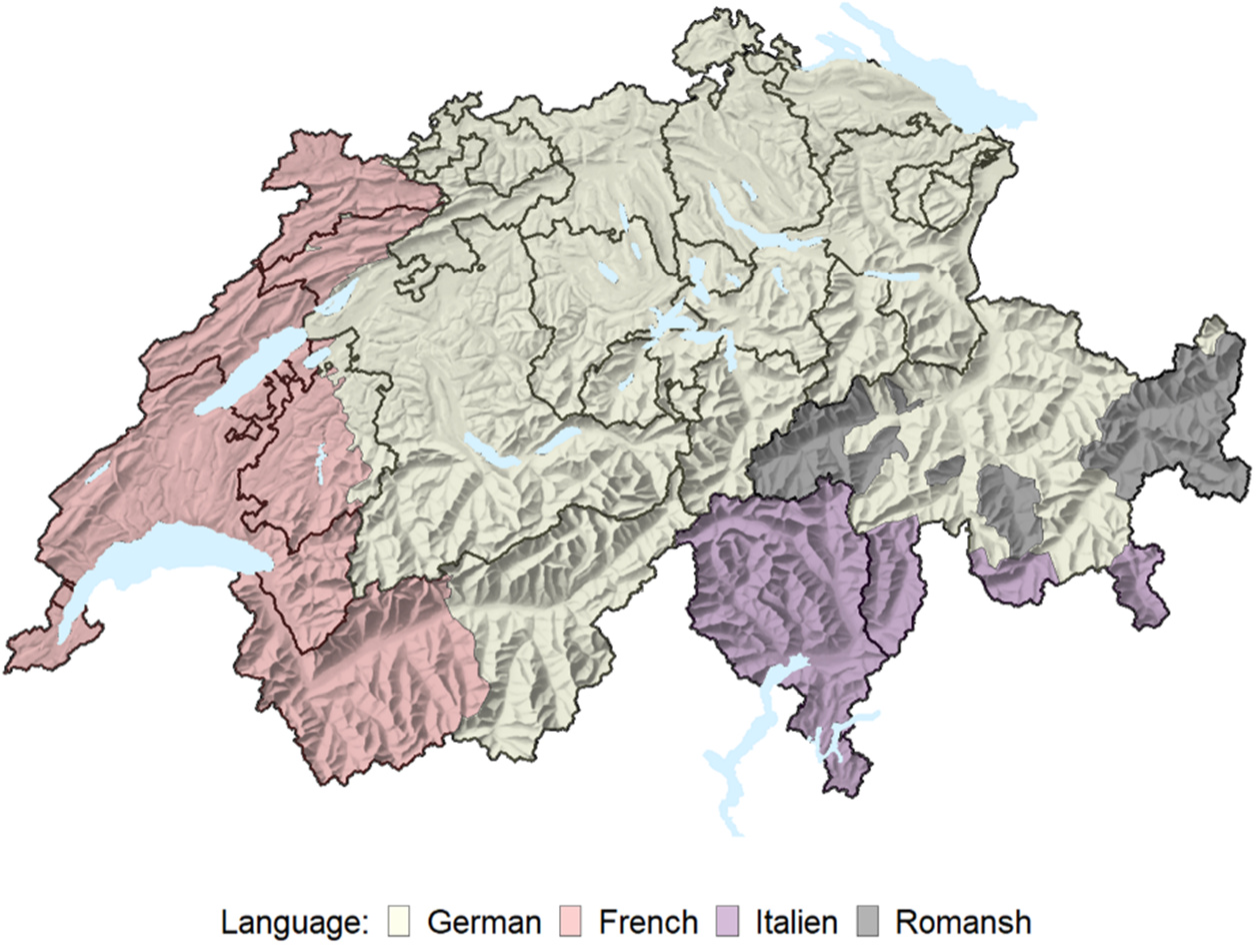
Main language region of Switzerland. Romanish is added to the German language region in the analysis.

**Supplement Figure 2:**
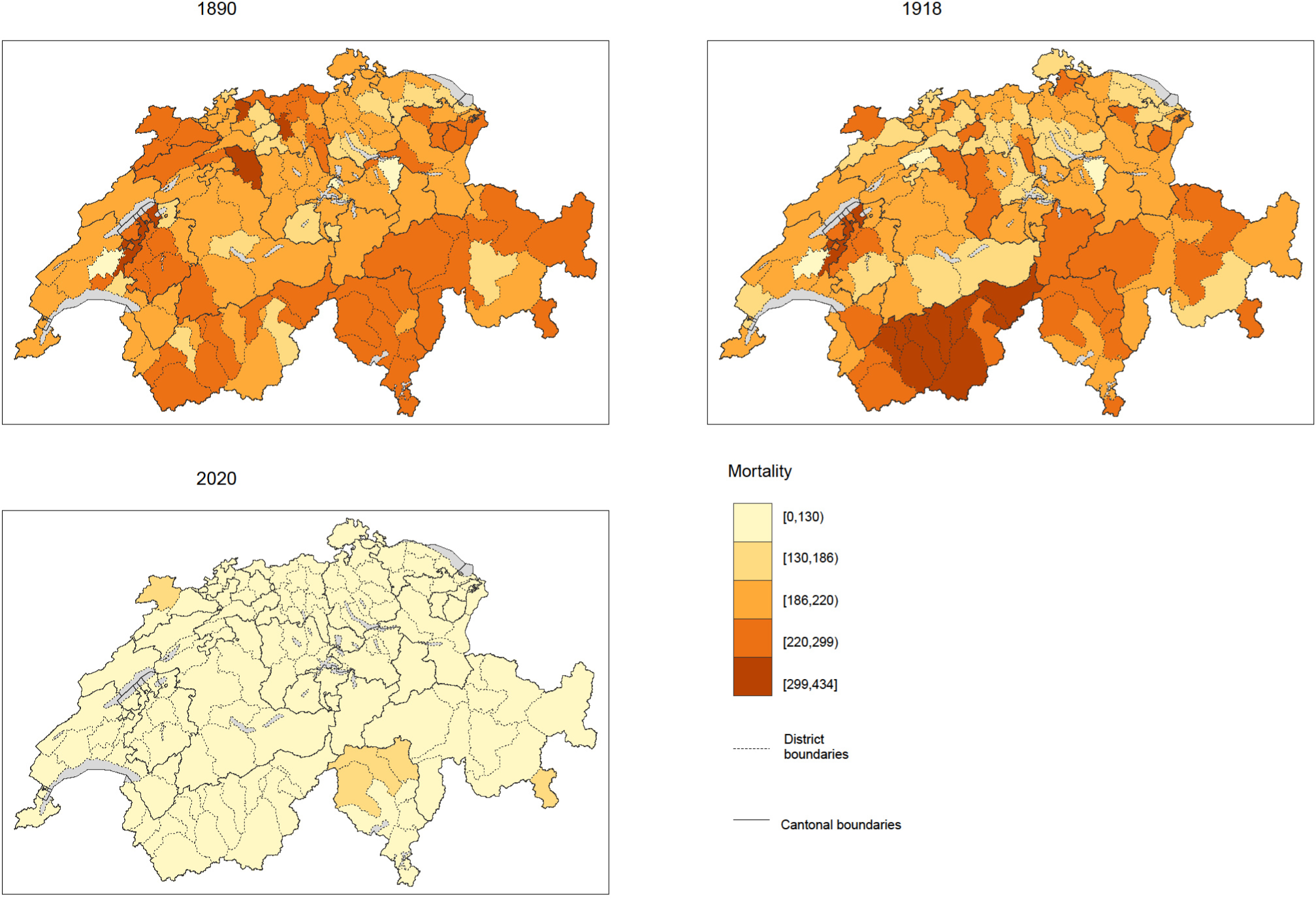
Mortality per 10’000 inhabitants.

**Supplement Figure 3:**
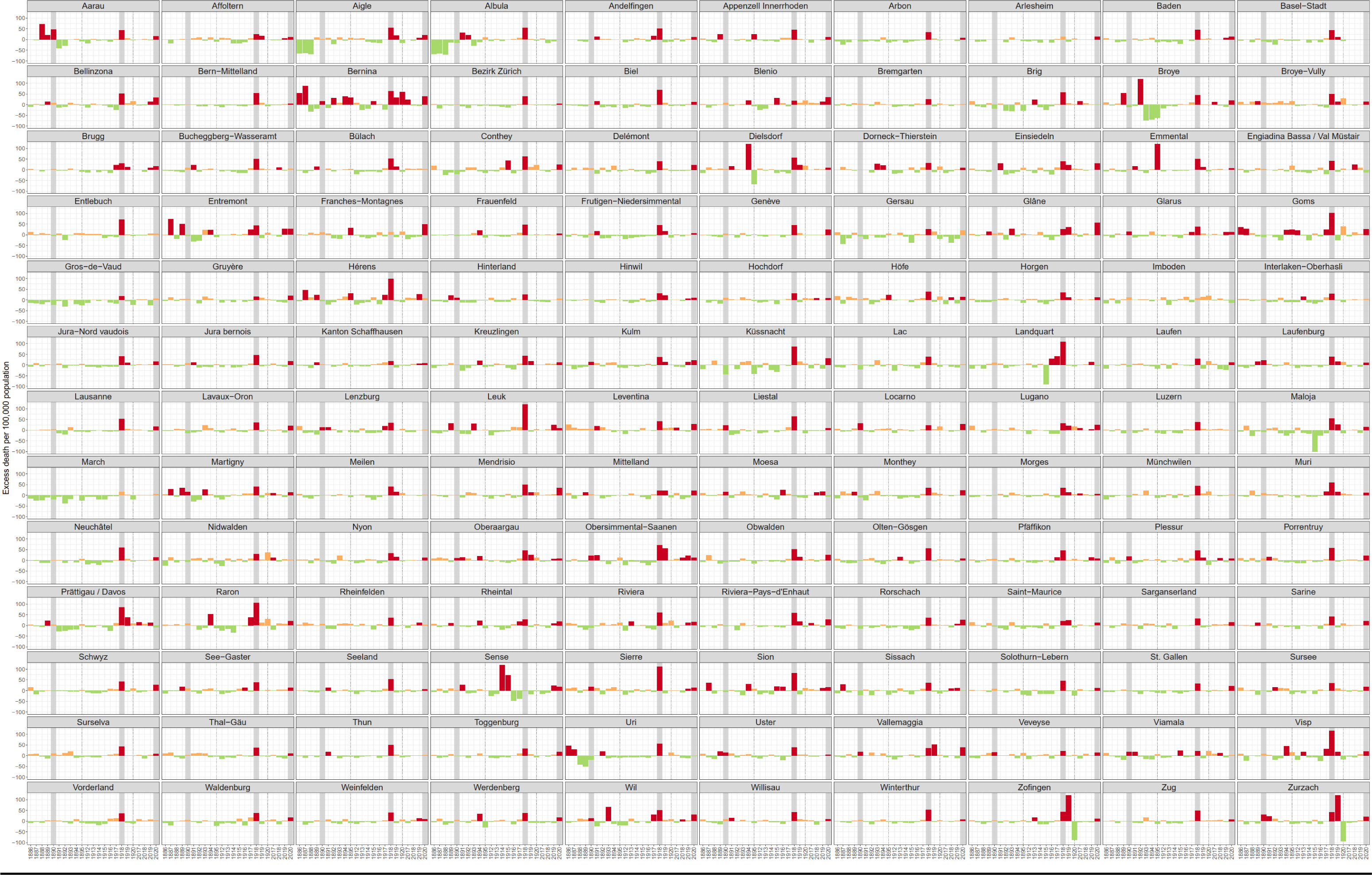
Excess mortality in percent for each district and the years 1886-1895, 1912-1920 and 2017-2020. Red bars indicate excess mortality.

**Supplement Figure 4:**
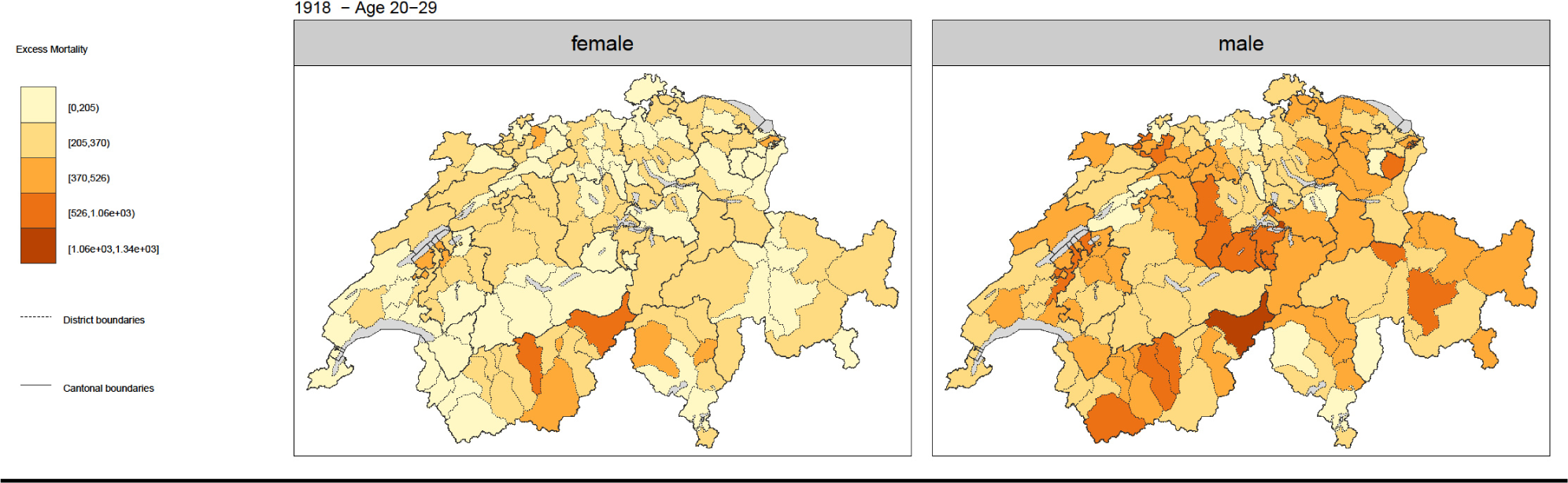
Excess mortality of the 1918, age 20-29 and sex.

**Supplement Table 1:**
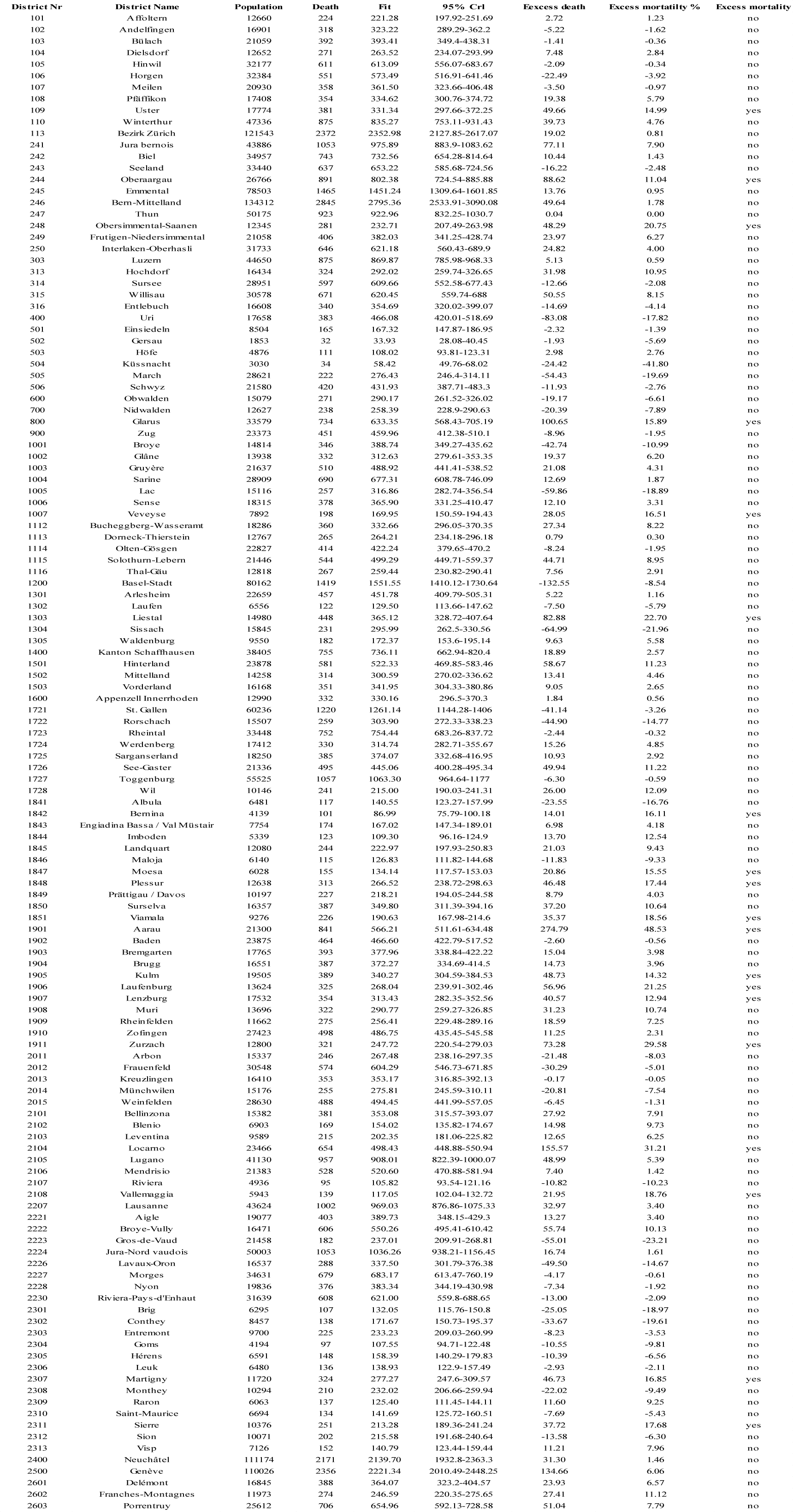
1890 excess mortality of each district.

**Supplement Table 2:**
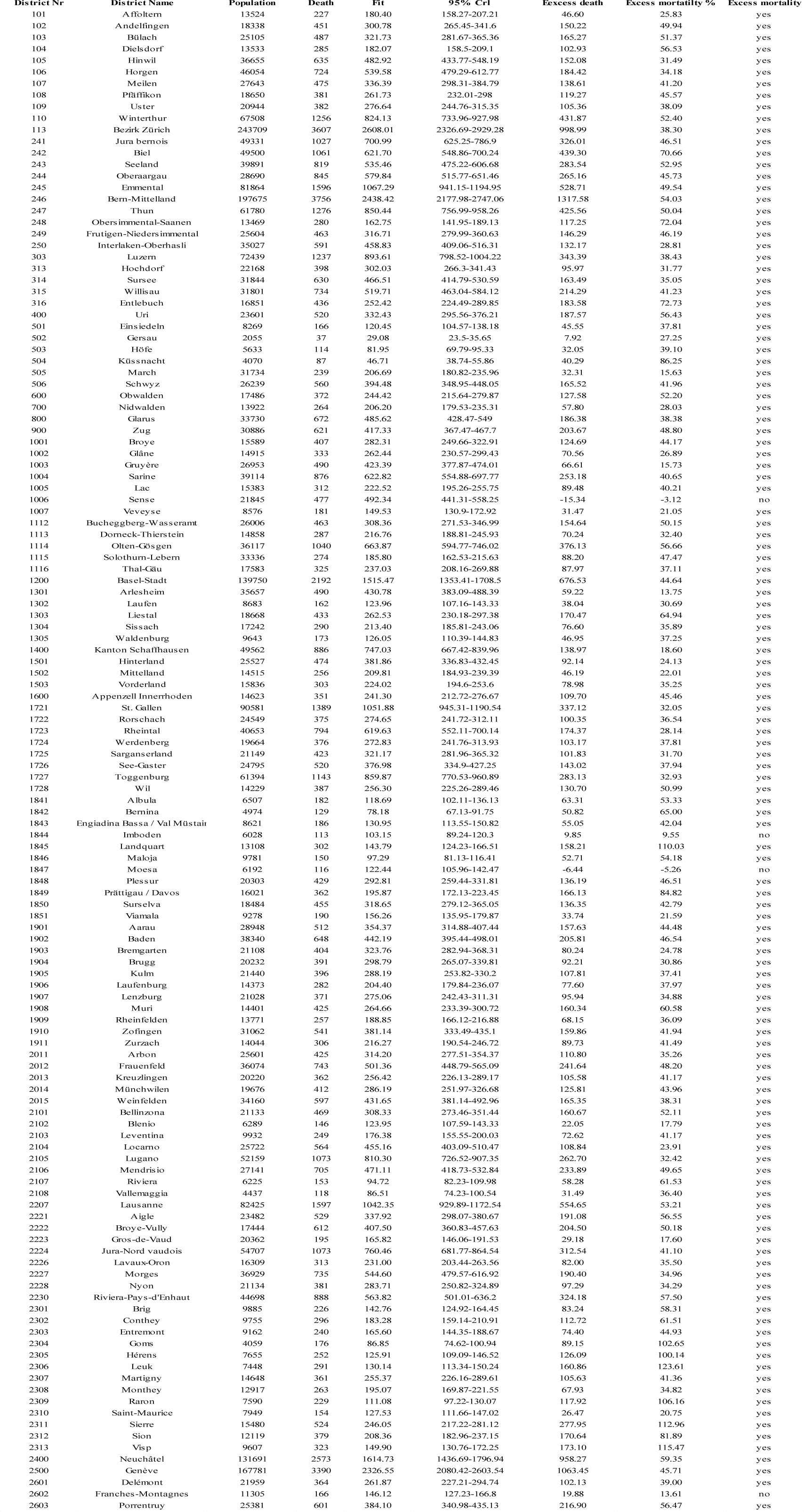
1918 excess mortality of each district.

**Supplement Table 3:**
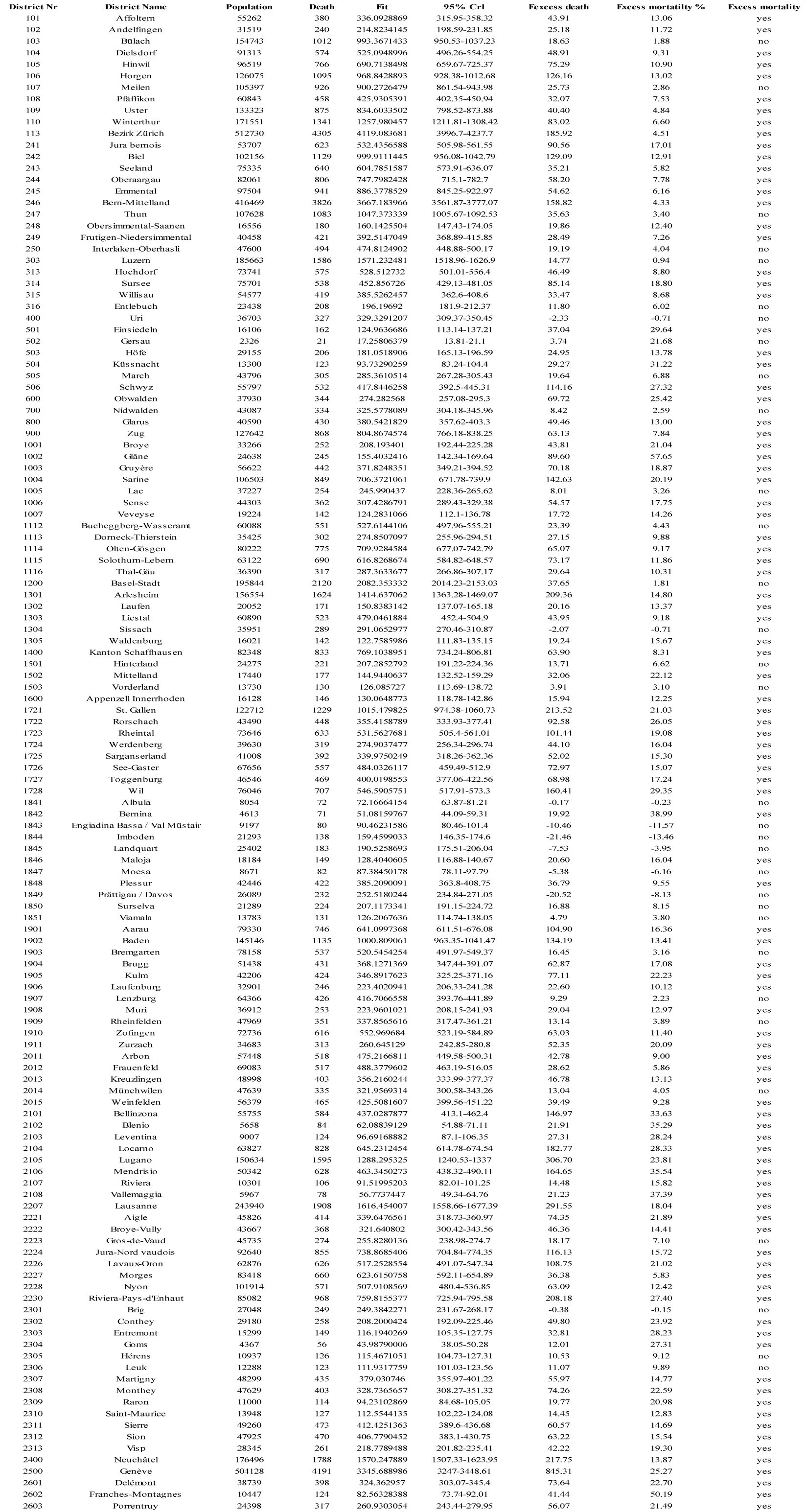
2020 excess mortality of each district.

**Supplement Figure 5:**
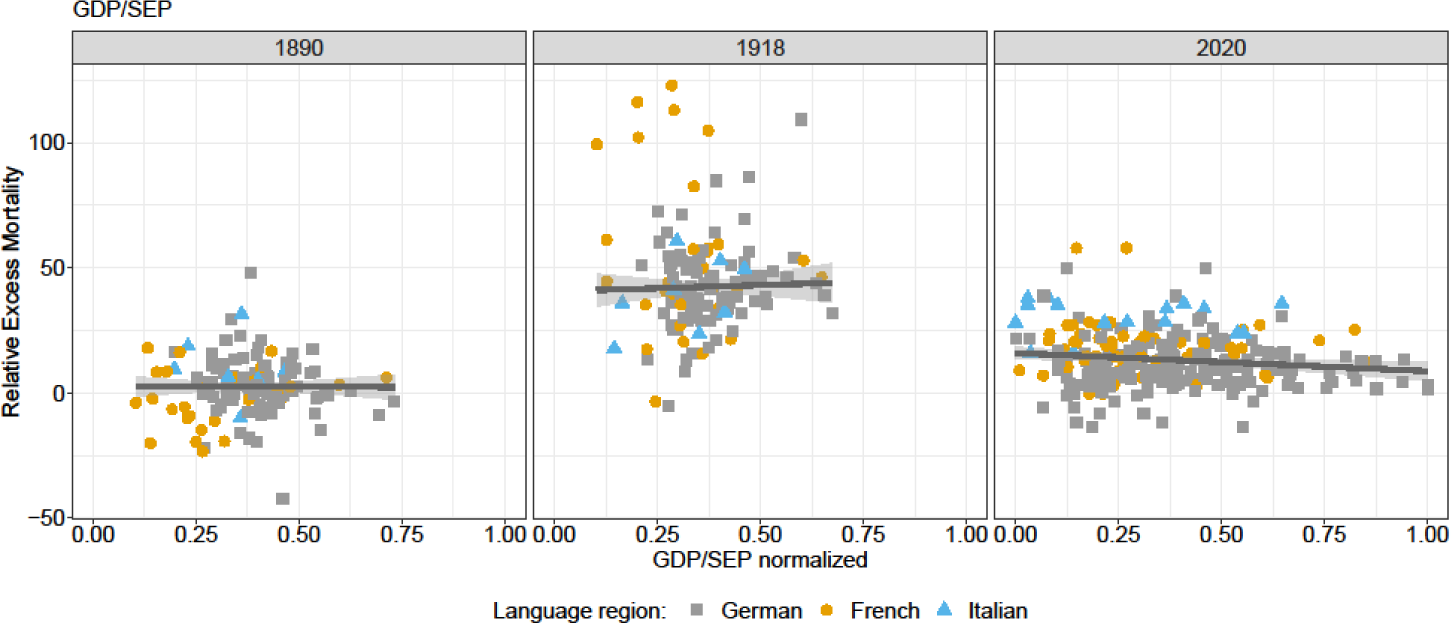
Robust linear regression of normalized GDP (1890 and 1918)/ SEP (2020) and relative excess mortality.

**Supplement Figure 6:**
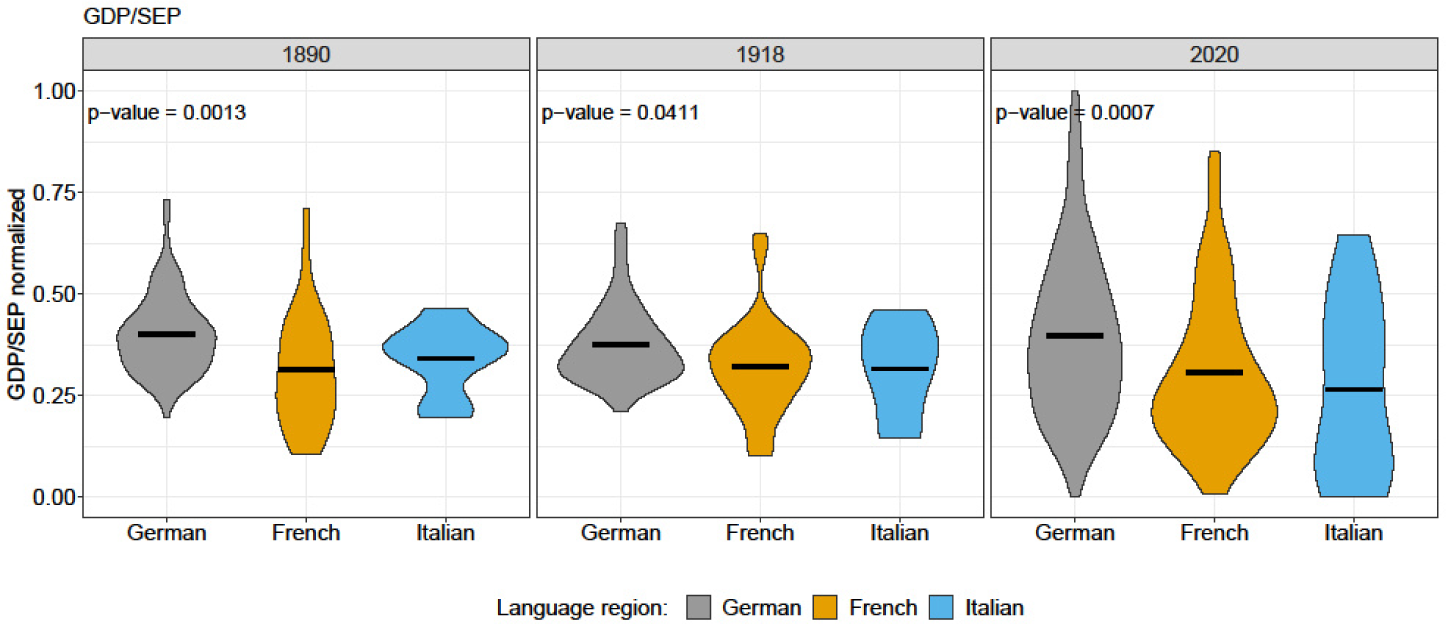
Violin plots of normalized GDP/SEP and results of Kruskal-Wallis Rank Sum Test between language regions for each year.

**Supplement Figure 7:**
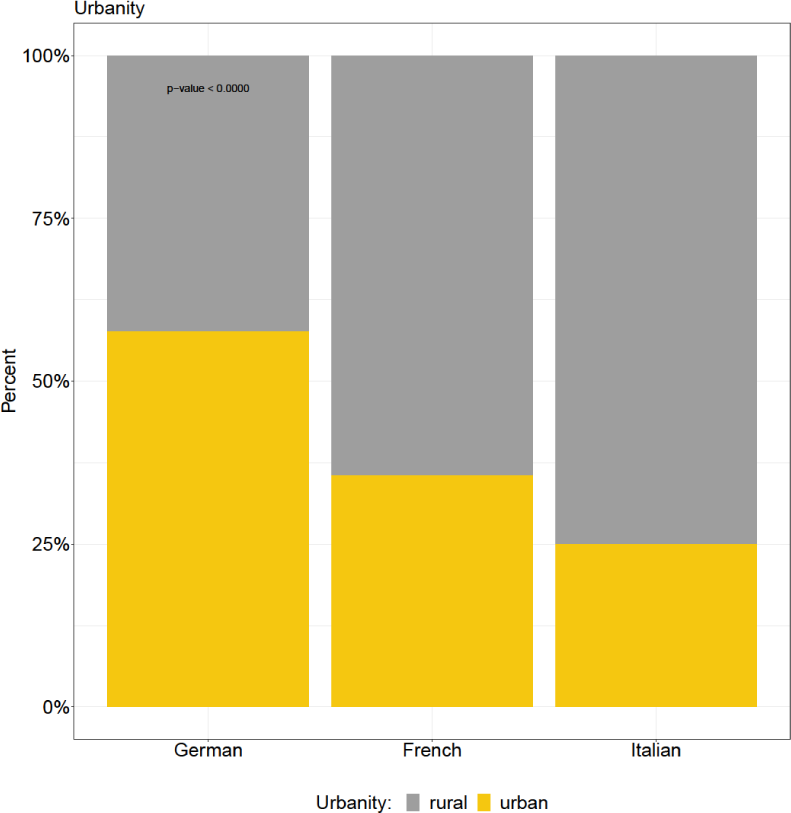
Barplot of urbanity for each language region and results of chi-square test.

**Supplement Figure 8:**
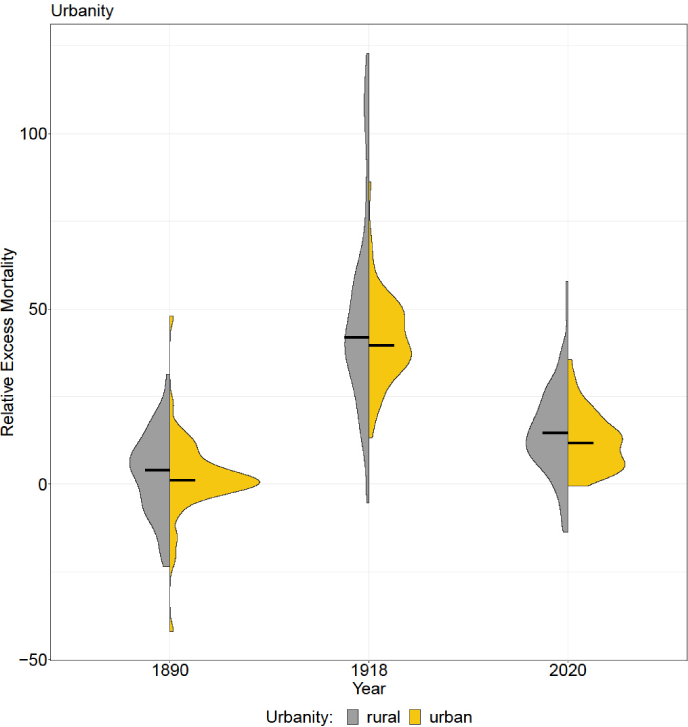
Violin plots of urbanity.

**Supplement Figure 9:**
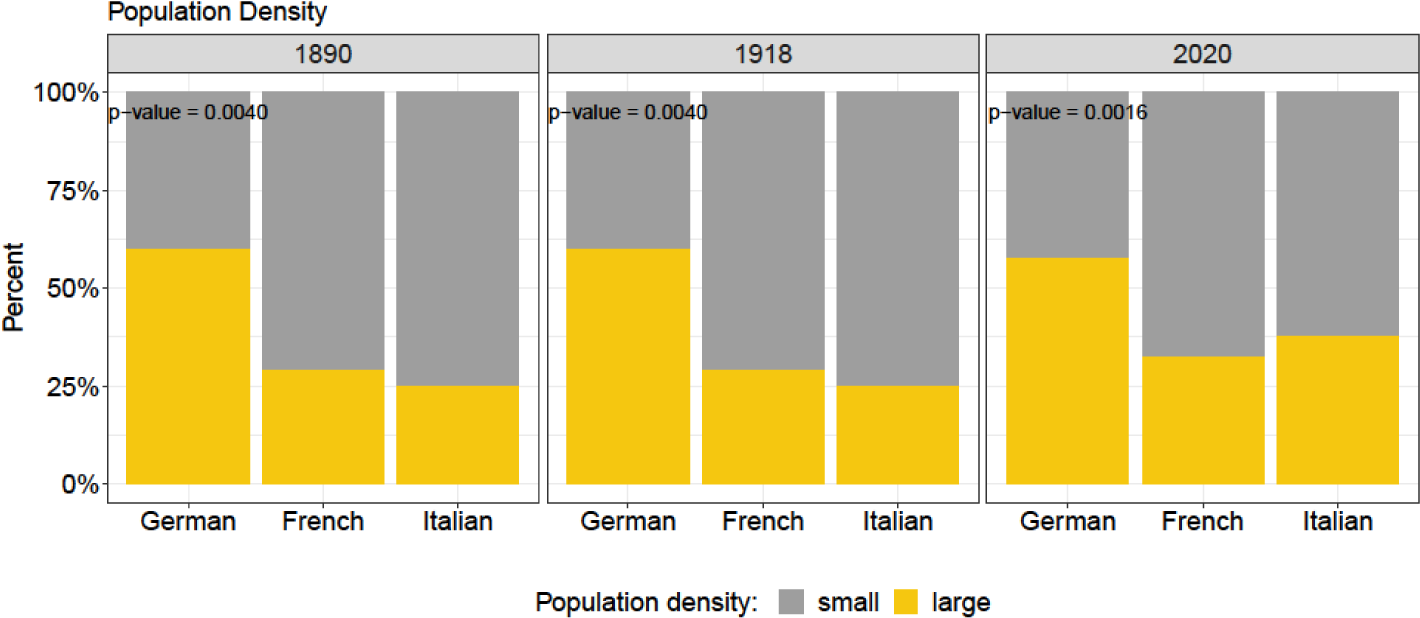
Barplot of population density for each language region and year and results of chi-square test.

**Supplement Figure 10:**
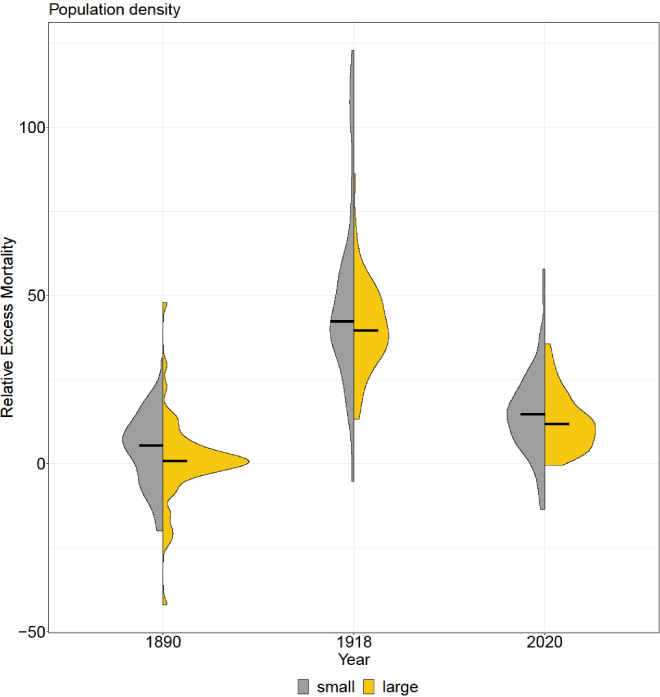
Violin plots of population density.

**Supplement Figure 11:**
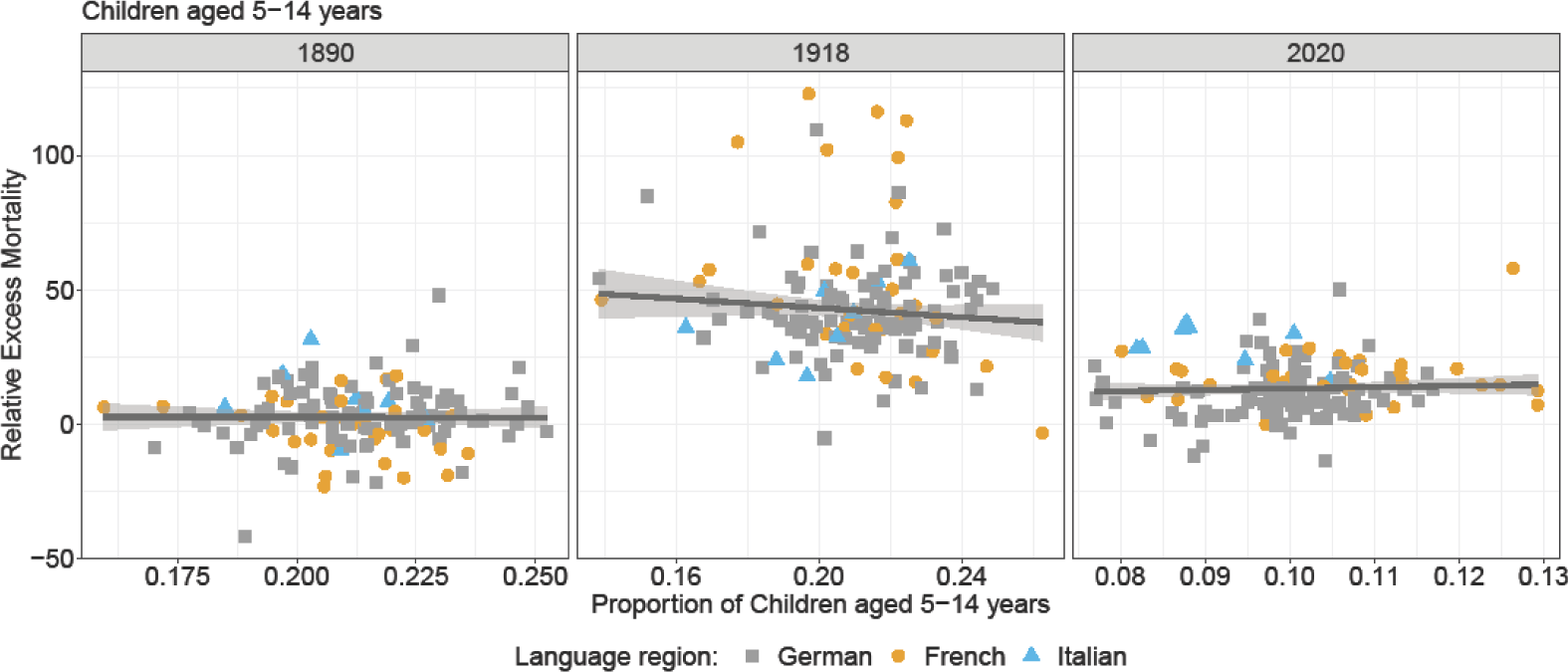
Robust linear regression of proportion of children 5-14 years and relative excess mortality.

**Supplement Figure 12:**
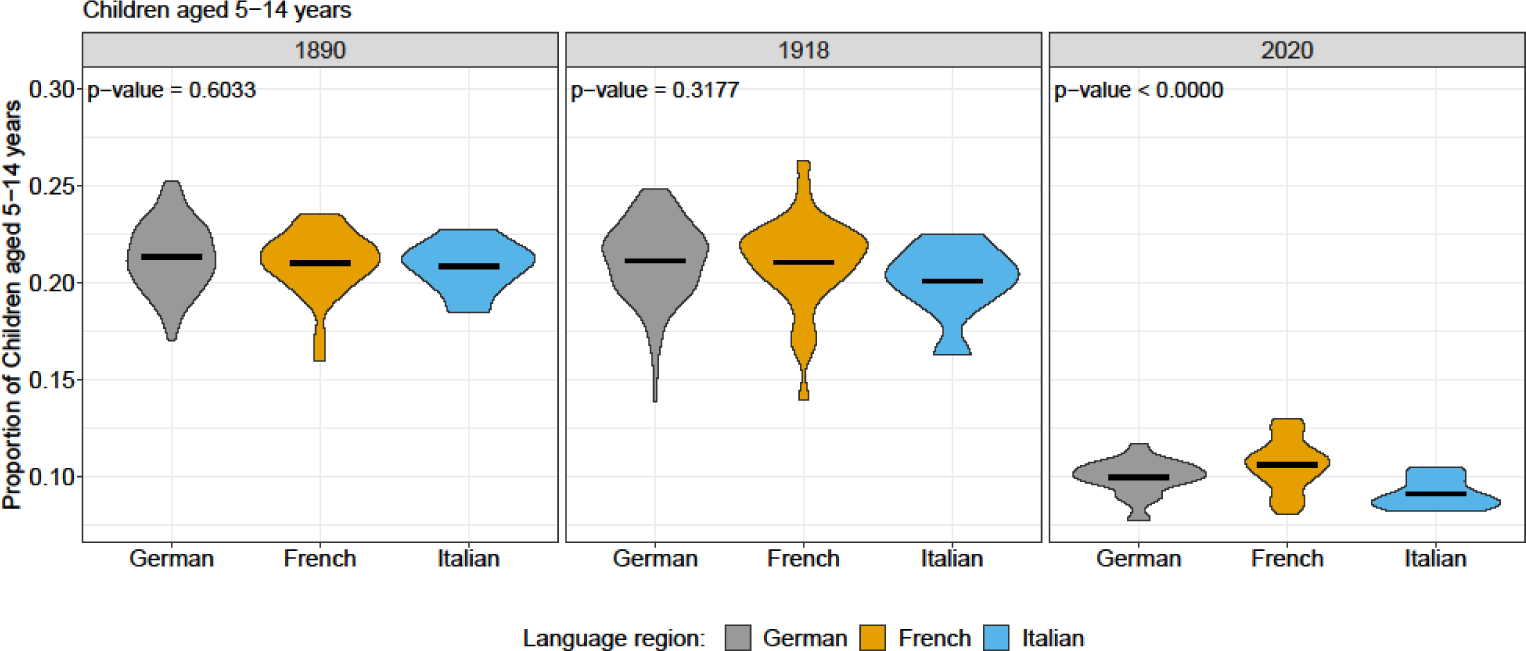
Violin plots of proportion of children 5-14 years old and results of Kruskal-Wallis Rank Sum Test between language regions for each year.

**Supplement Figure 13:**
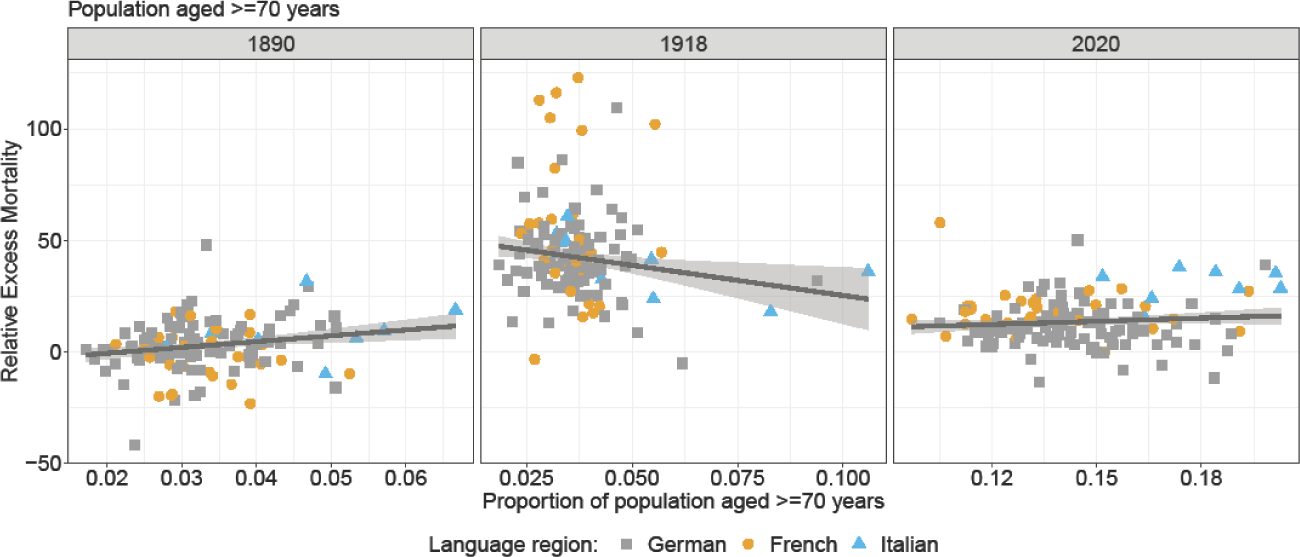
Robust linear regression of proportion of ≥ 70 years old and relative excess mortality.

**Supplement Figure 14:**
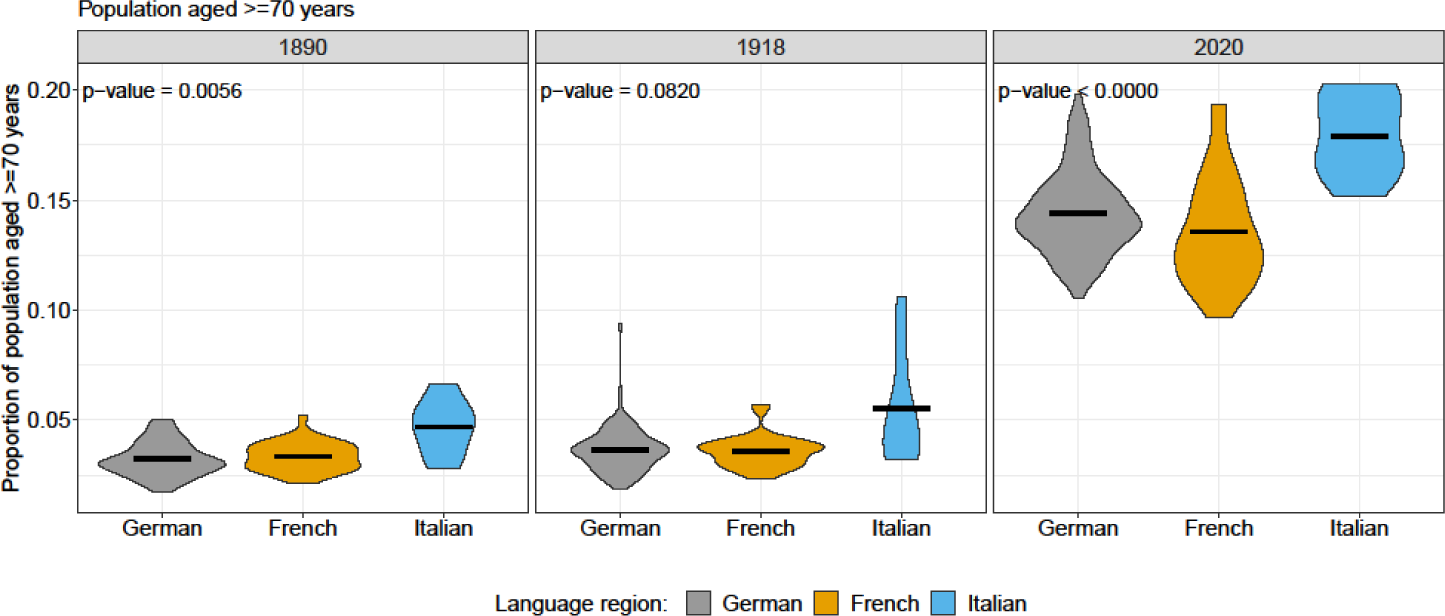
Violin plots of proportion of ≥ 70 years old and results of Kruskal-Wallis Rank Sum Test between language regions for each year.

**Supplement Figure 15:**
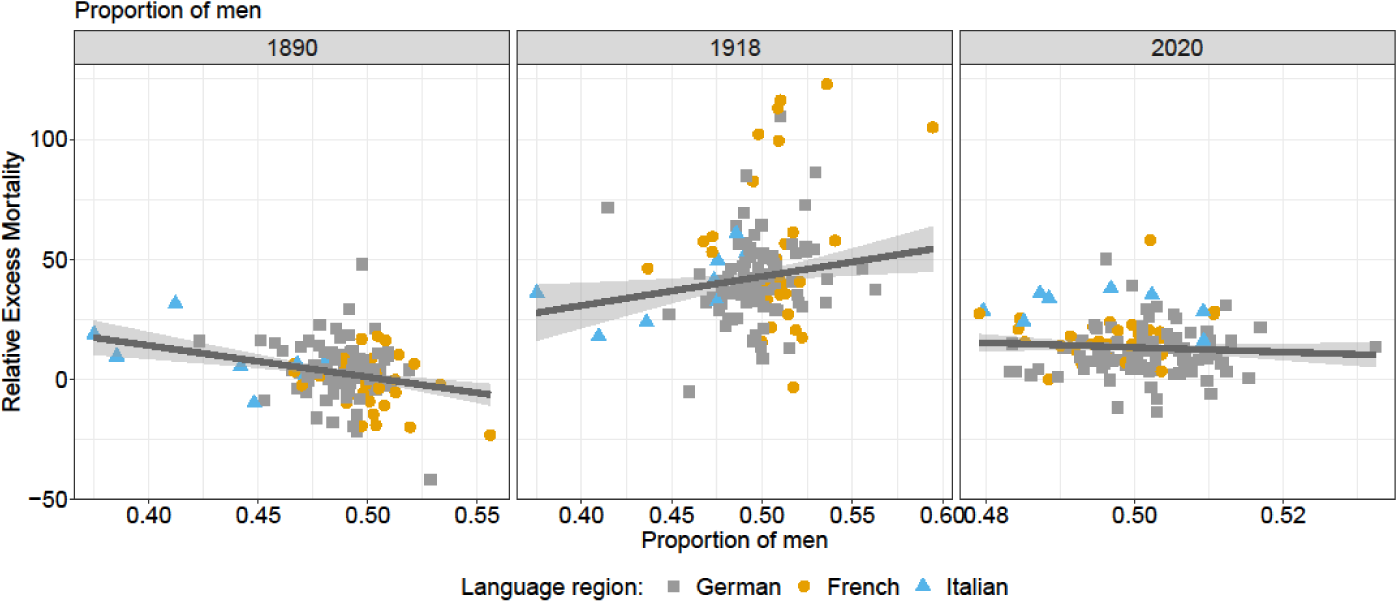
Robust linear regression of proportion of men and relative excess mortality.

**Supplement Figure 16:**
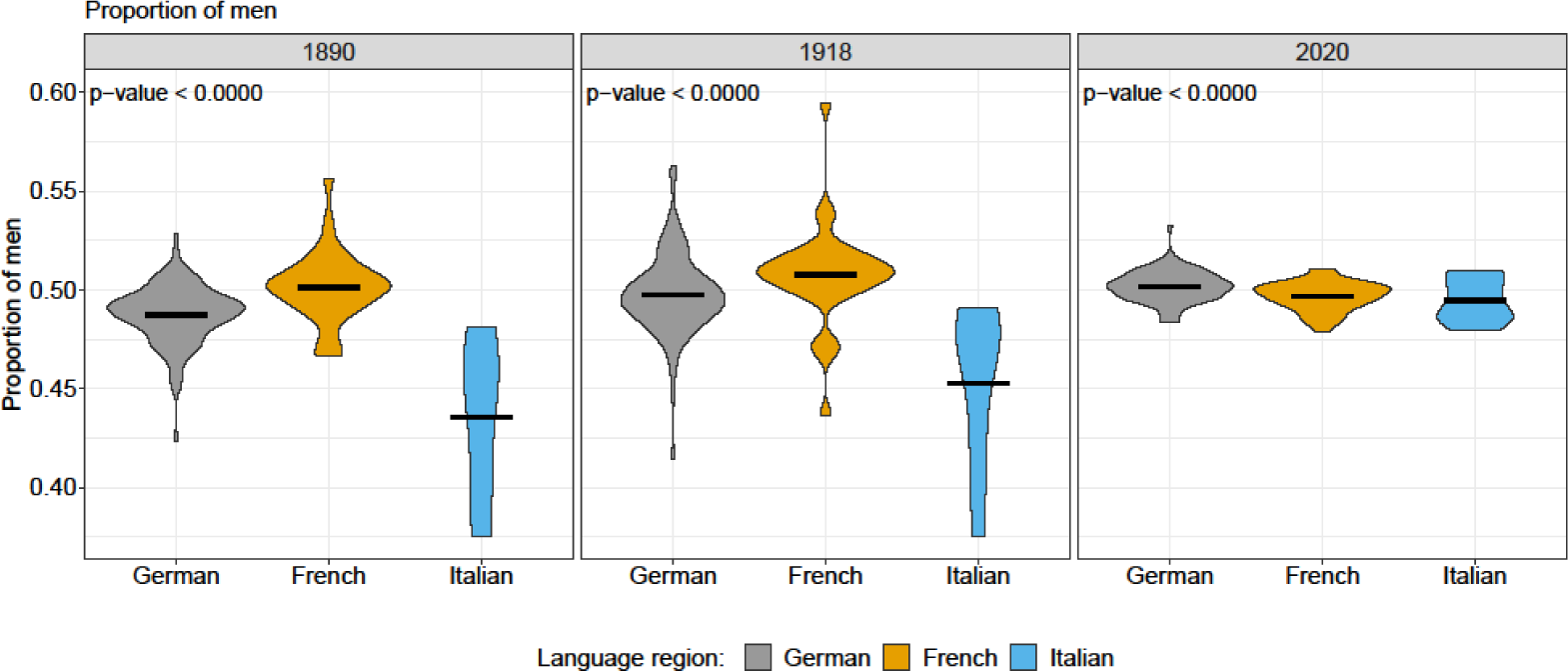
Violin plots of proportion of men and results of Kruskal-Wallis Rank Sum Test between language regions for each year.

**Supplement Figure 17:**
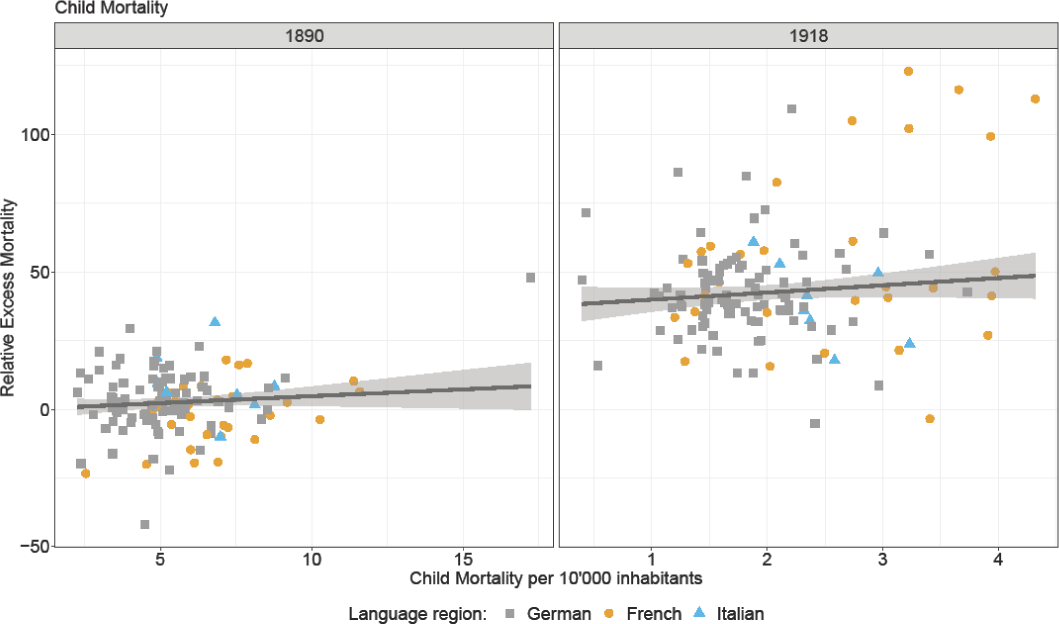
Robust linear regression of child mortality and relative excess mortality.

**Supplement Figure 18:**
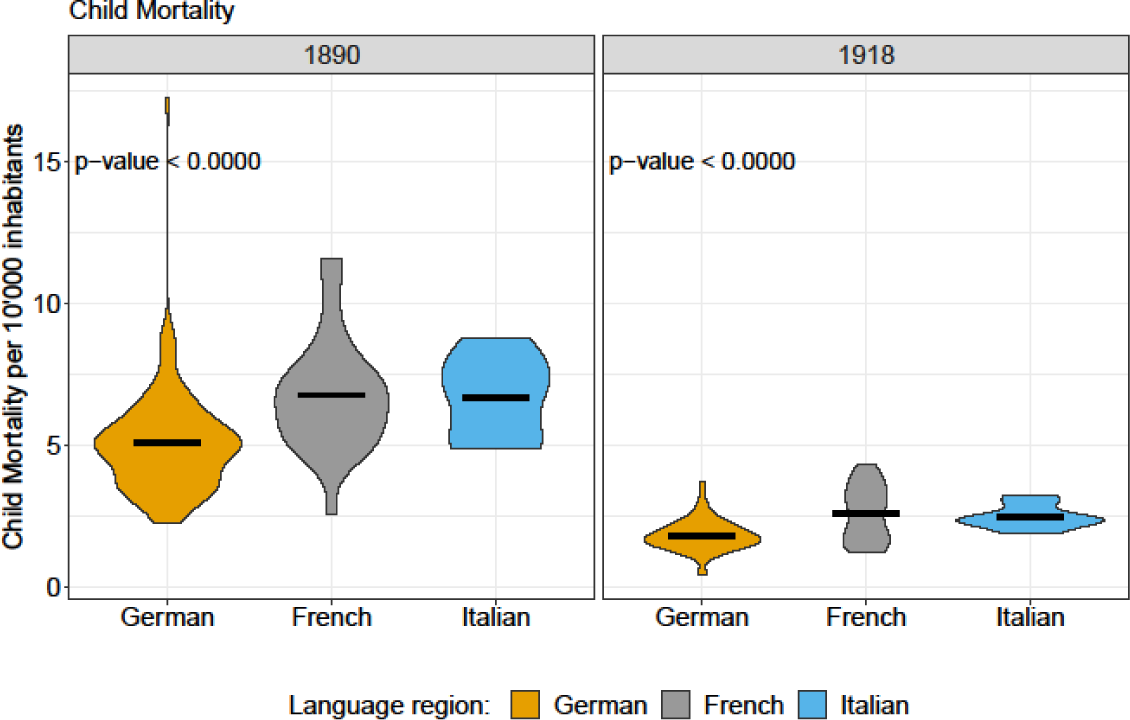
Violin plots of proportion of child mortality and results of Kruskal-Wallis Rank Sum Test between language regions for each year.

**Supplement Figure 19:**
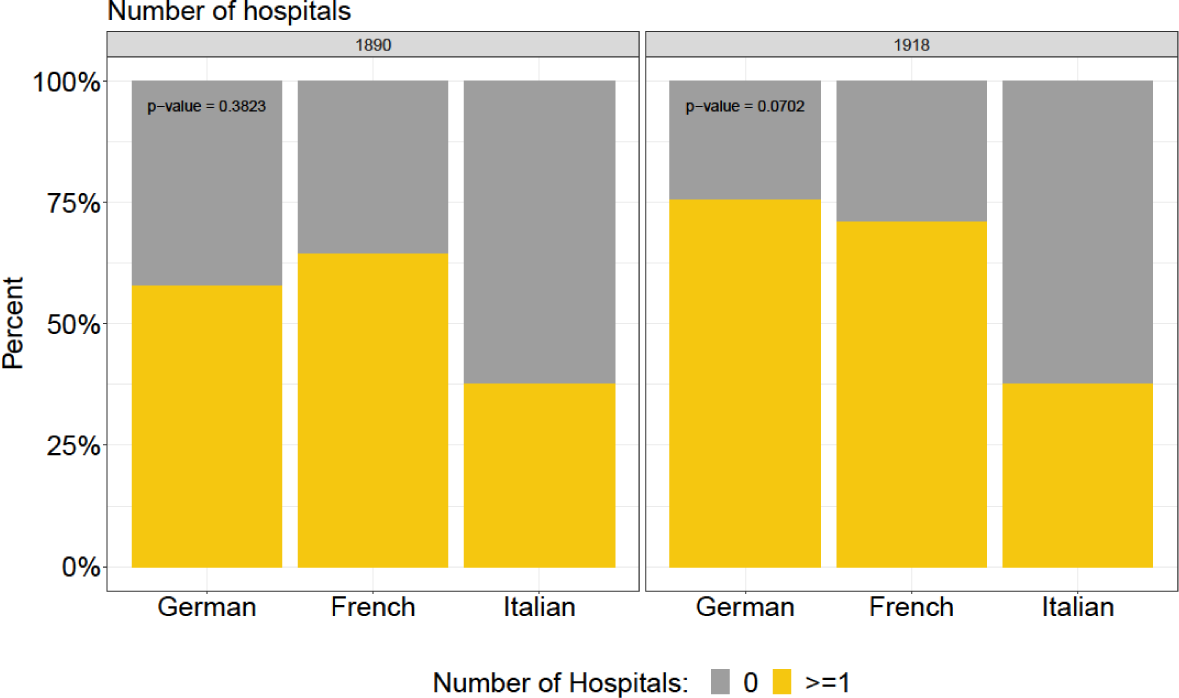
Barplot of hospitals for each language region and year and results of chi-square test.

**Supplement Figure 20:**
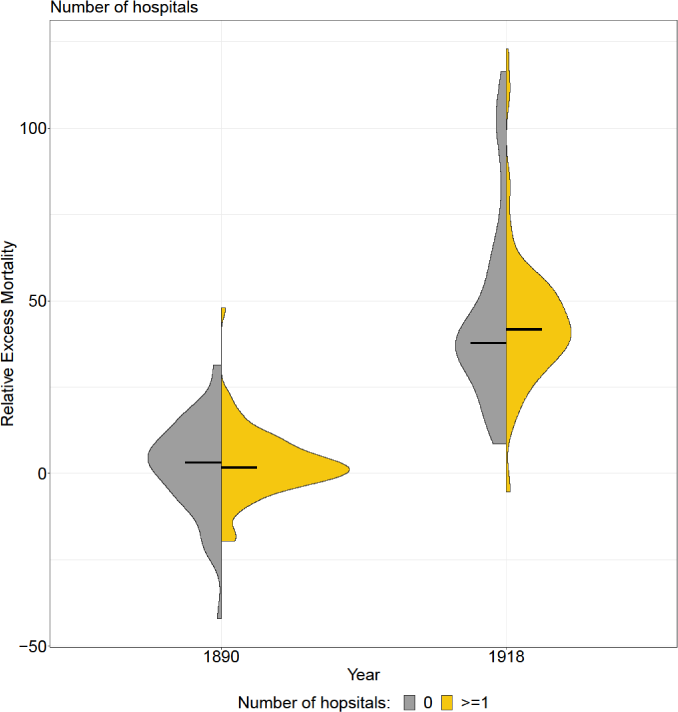
Violin plots of hospitals.

**Supplement Figure 21:**
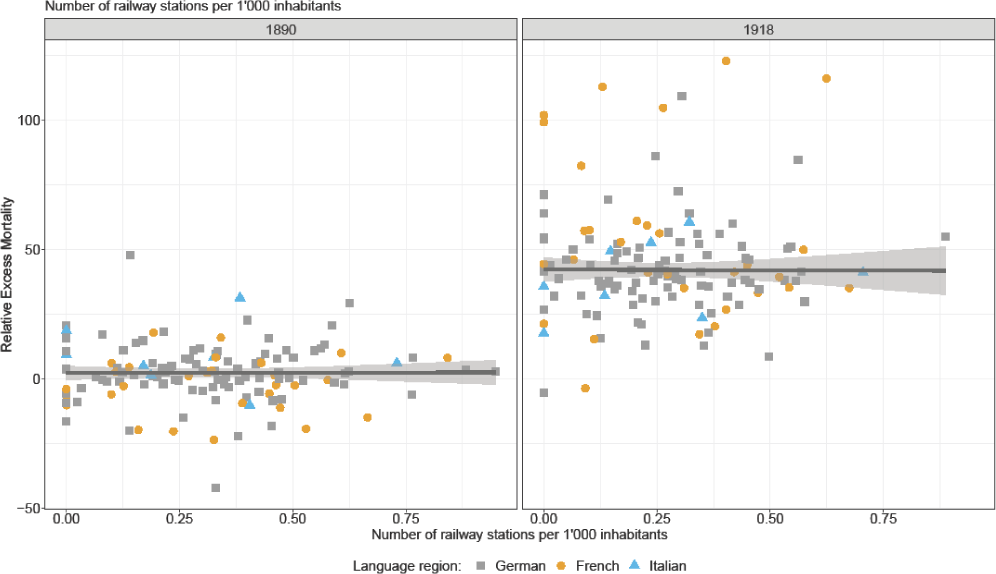
Robust linear regression of railway stations of 1’000 inhabitants and relative excess mortality.

**Supplement Figure 22:**
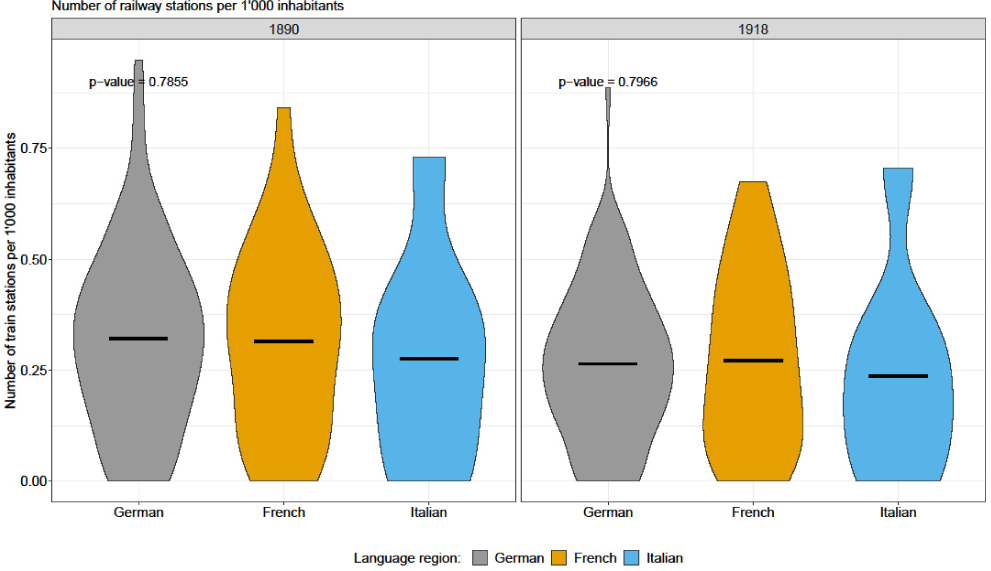
Violin plots of proportion of railway stations of 1’000 inhabitants and results of Kruskal-Wallis Rank Sum Test between language regions for each year.

**Supplement Figure 23:**
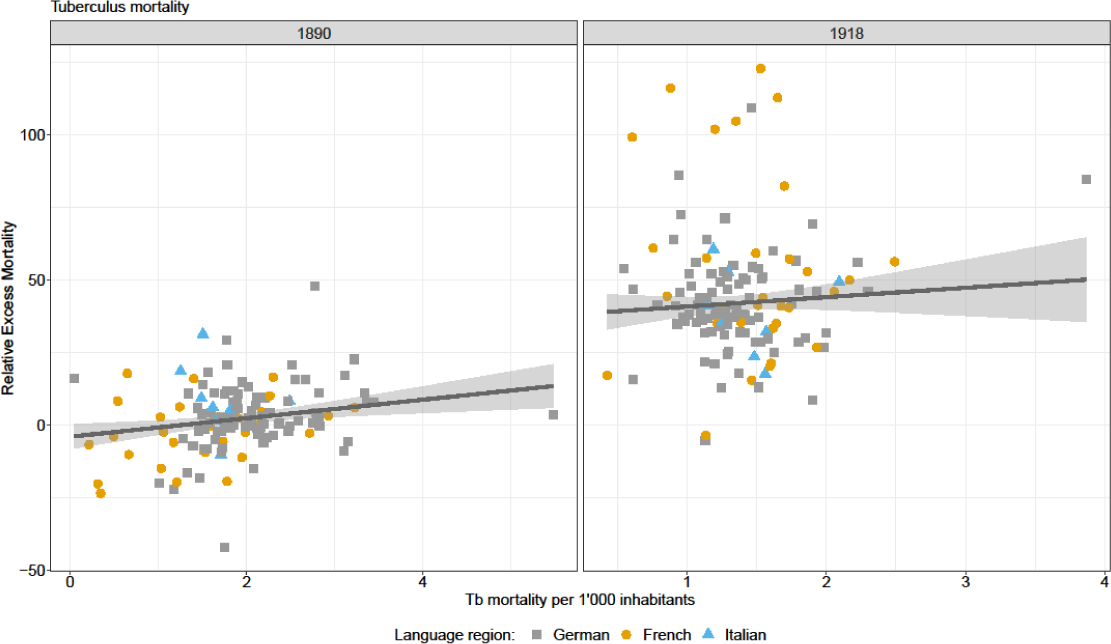
Robust linear regression of TB mortality and relative excess mortality.

**Supplement Figure 24:**
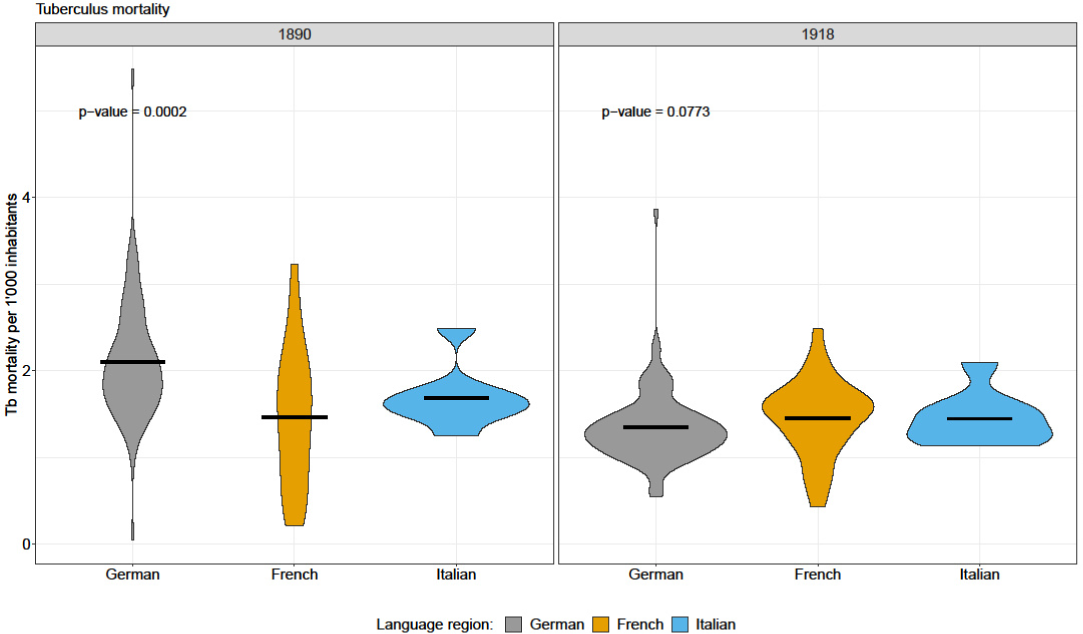
Violin plots of TB mortality and results of Kruskal-Wallis Rank Sum Test between language regions for each year.

**Supplement Table 4:** Coefficient and 95% confidence interval of univariate robust linear regression with excess mortaliy.

| **Cofactor** | **Year** | **Coefficient (95% CI)** |
| --- | --- | --- |
| GDP / SEP | 1890 | -0.42 (-16.02 to 15.17) |
| GDP / SEP | 1918 | 5.06 (-21.21 to 31.33) |
| GDP / SEP | 2020 | -11.75 (-19.57 to -3.93) |
| Urbanicity (Ref: Rural) | 1890 | -1.21 (-4.88 to 2.47) |
| Urbanicity (Ref: Rural) | 1918 | -3.16 (-8.53 to 2.21) |
| Urbanicity (Ref: Rural) | 2020 | -2.95 (-6.41 to 0.51) |
| Large population density (Ref: Small) | 1890 | -3.64 (-7.23 to -0.06) |
| Large population density (Ref: Small) | 1918 | -3.92 (-9.38 to 1.53) |
| Large population density (Ref: Small) | 2020 | -2.19 (-5.64 to 1.26) |
| Proportion of 5-14 year-olds | 1890 | -0.01 (-1.06 to 1.03) |
| Proportion of 5-14 year-olds | 1918 | -0.91 (-2.23 to 0.41) |
| Proportion of 5-14 year-olds | 2020 | 0.51 (-1.15 to 2.18) |
| Proportion of >=70 year-olds | 1890 | 2.63 (0.66 to 4.6) |
| Proportion of >=70 year-olds | 1918 | -2.59 (-4.83 to -0.36) |
| Proportion of >=70 year-olds | 2020 | 0.48 (-0.32 to 1.28) |
| Proportion of men | 1890 | -24.12 (-37.02 to -11.22) |
| Proportion of men | 1918 | 25.54 (3.07 to 48) |
| Proportion of men | 2020 | -5.23 (-16.45 to 5.99) |
| Proportion of child mortality | 1918 | 2.39 (-1.35 to 6.12) |
| Train station per inhabitant | 1890 | 0.19 (-8.32 to 8.7) |
| Train station per inhabitant | 1918 | -0.97 (-16.32 to 14.37) |
| Tuberculosis mortality | 1890 | 3.24 (0.93 to 5.54) |
| Tuberculosis mortality | 1918 | 2.84 (-3.67 to 9.34) |

